# CombiANT Reader - Deep learning-based automatic image processing and measurement of distances to robustly quantify antibiotic interactions

**DOI:** 10.1101/2024.10.16.24315598

**Authors:** Erik Hallström, Nikos Fatsis-Kavalopoulos, Manos Bimpis, Anders Hast, Dan I. Andersson

## Abstract

Antibiotic resistance is a severe danger to human health, and combination therapy with several antibiotics has emerged as a viable treatment option for multi-resistant strains. CombiANT is a recently developed agar plate-based assay where three reservoirs on the bottom of the plate create a diffusion landscape of three antibiotics that allows testing of the efficiency of antibiotic combinations. This test, however, requires manually assigning nine reference points to each plate, which can be prone to errors, especially when plates need to be graded in large batches and by different users. In this study, an automated deep learning-based image processing method is presented that can accurately segment bacterial growth and measure more than 150 distances from key points on the CombiAnt assay at sub-millimeter precision. The software was tested on 100 plates using photos captured by three different users with their mobile phone cameras, comparing the automated analysis with the human scoring. The result indicates significant agreement between the users and the software. Moreover, the automated analysis remains consistent when applied to different photos of the same assay despite varying photo qualities and lighting conditions. The software can easily be integrated into a potential smartphone application. Integrating deep learning-based smartphone image analysis with simple agar-based tests like CombiANT could unlock powerful tools for combating antibiotic resistance.

**Author Summary:** Antibiotic resistance is a significant problem worldwide with increasing prevalence of multi-resistant bacteria that may require the simultaneous administration of several different antibiotics. With the right antibiotics and concentration, such combination therapy may treat a strain that is otherwise resistant to each antibiotic individually. CombiANT is a novel test that can be used to identify suitable or inappropriate antibiotic combinations. However, it requires the human evaluator to grade each plate manually, which is time-consuming, and errors can easily be made, especially if the human evaluator needs to grade many plates in succession. In this study, an image processing pipeline is developed using a deep neural network to grade CombiANT test assays automatically.

## 1 Introduction

Antibiotics are a cornerstone of modern healthcare, allowing treatment of bacterial infections that were once fatal or severely disabling. However, their effectiveness is increasingly threatened by antibiotic resistance, driven by antibiotic overuse or misuse [1]. One way to combat the resistance is by prescribing combination therapies.

Some antibiotics can work together in synergy, resulting in a greater efficacy than when each one acts alone. Conversely, some antibiotics may neutralize each other’s effects, a phenomenon referred to as antagonistic effects [2] [3] [4] [5] [6]. This interaction is also dependent on the respective concentration of each antibiotic. Therefore, it is of high importance to carefully evaluate and understand the interactions between different antibiotics before prescribing combination therapies.

One novel method to assess antibiotic interactions is CombiANT, an easy-to-use assay that allows for the testing of antibiotic synergies on agar plates (Fatsis-Kavalopoulos et al.) [7]. The CombiANT test involves an agar plate containing three 3D-printed reservoirs, each accommodating a different antibiotic. Following preparation, a diffusion landscape forms, generating different concentrations of each antibiotic at every location on the plate. The assay is shown in Fig 1 with the inserts containing antibiotics marked with A, B, and C. Opposite of each reservoir, at the outer side of the white circle mark, the antibiotic acts alone as the concentration of the others there is negligible. The inscribed white triangle mark constitutes the interaction area, where antibiotics act in pairs, with the highest combination concentration at the triangle vertices. Due to the distance of the opposite reservoir, the concentration of the third antibiotic is negligible at each triangle vertex. Antibiotics A and B act together in the bottom right, A and C in the bottom left, and B and C in the top of the triangle. The darker areas in the assay exhibit uninhibited bacterial growth. The assay has two growth zones: an ”inner” inside the interaction zone triangle and an ”outer” outside the circle. In the original CombiANT test, a human has to manually annotate key points on the edges of the growth zones and also pinpoint the triangle vertices: ICA, ICB, and ICC are inhibitory concentration points placed at the bacteria boundary opposite the midpoint of the corresponding reservoir on a straight line perpendicular to the circle perimeter. Three combination inhibitory points, CPAB, CPAC, and CPBC, are placed on the rim of the inner growth zone at the closest point to the respective triangle vertex. Finally, the evaluator has to annotate the vertices of the triangle mark interaction zone in the correct order: VAB, VAC, VBC, enabling the original CombiAnt software to align all coordinates on the assay with the pre-calculated diffusion landscape. After this manual point-annotation, the required distances can be obtained, shown as dashed lines in Fig 1.

**Fig 1.**
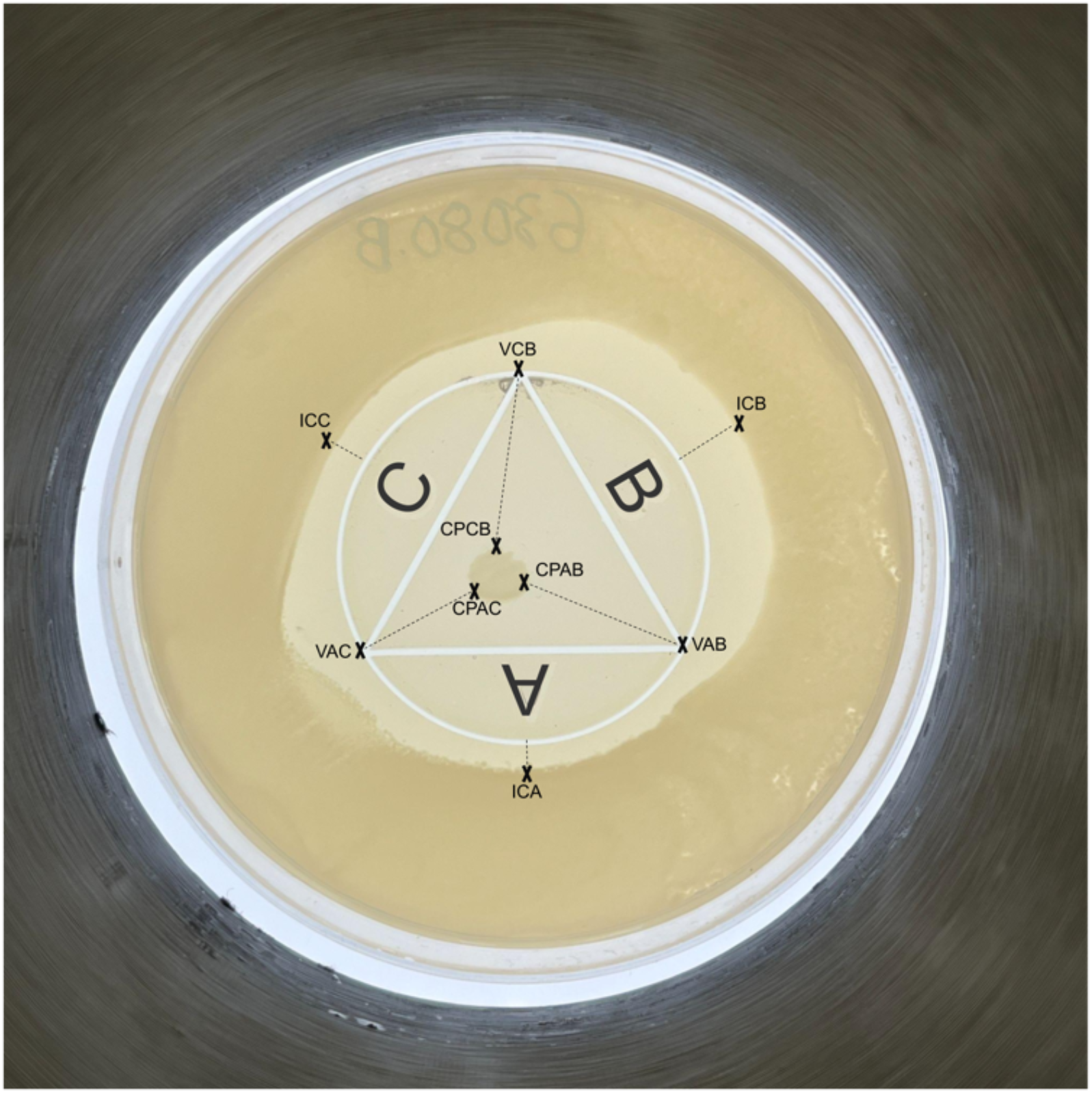
CombiANT. A CombiANT assay with nine annotated key points required to be manually pinpointed by a human evaluator. Dashed lines outline the obtained distances. ICA, ICB, and ICC are inhibitory concentration points that indicate the effects of the corresponding antibiotics acting alone. Due to irregularities of the outer growth zone boundary, this value can be inconsistent. CPAB, CPAC, and CPBC are combination inhibitory points, and VAC, VAB, and VABC are triangle vertices. The three antibiotic inserts (A, B, and C) are filled in black for visibility.

At present no computer vision application for automatically scoring the CombiAnt assay is available, even though some related image processing methods exist, including the automated counting of colony-forming units (CFU) on an agar plate [8]. This task relates to the growth-zone segmentation in the CombiANT assay in the proposed pipeline, as both require identifying and segmenting bacterial content on the agar plate while discarding non-bacterial objects and artifacts. Several image processing methods have been proposed to accomplish this, utilizing both classical [9] [10] [11] [12] [13] [14] [15] and deep learning-based [16] [17] [18] [19] approaches. Another related image processing task is measuring the inhibition zone in the The Kirby–Bauer disk diffusion test. This zone is typically circular, hence, the Hough Circle transform is commonly used to measure the inhibition [20] [21], although various deep-learning models have been explored [22]. In the CombiANT test, the inhibition zone is not circular; hence, this prior can not be used.

To address this lack of automated analysis of antibiotic interactions, a deep-learning [23] based image processing pipeline was developed in this study for this specific task. The pipeline successfully scored all plates similar to the human evaluators, hence the overall aim of this study was achieved.

## 2 Results

The proposed CombiANT Reader software method automates the annotation process described in the original CombiANT test. The software finds the growth zones and triangle vertices, and measures the required distances at sub-millimeter precision, visualized in Fig 2. The main advantage of this automated approach compared to the existing CombiAnt methodology is reliability, as currently, the human evaluator has to annotate the points on the plate (shown in Fig 1) in the correct order.

**Fig 2.**
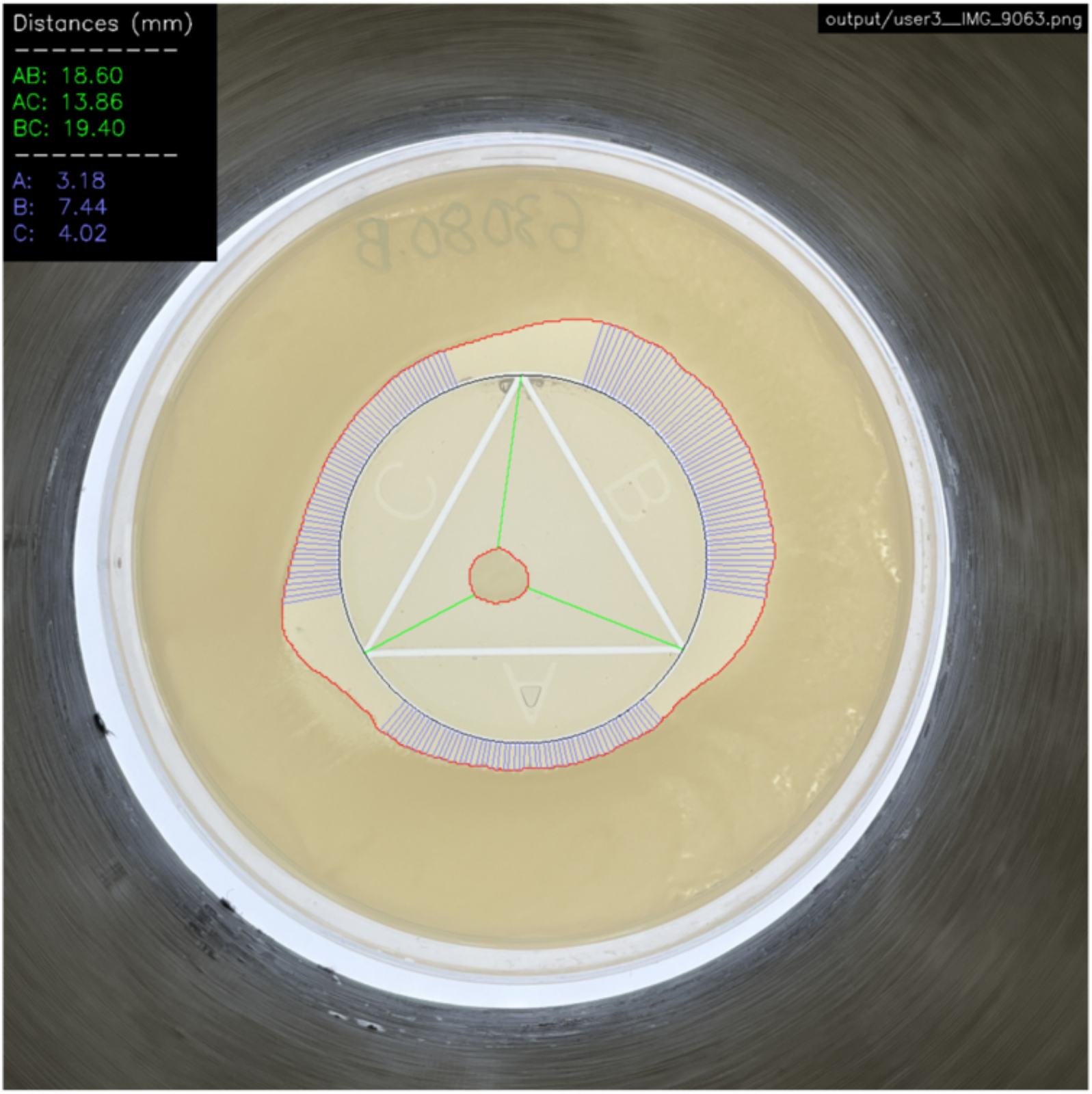
CombiANT Reader. The proposed CombiANT Reader software automatically finds the circle mark with the inscribed triangle, outlined with a black border, and the edges of the growth zones, outlined in red. The software measures 55 distances from each reservoir to the outer growth zone (blue lines) and automatically finds the closest distance to the inner growth zone from every triangle vertex (green lines). The legend shows the calculated distances in millimeters with two decimals. For the outer distances, the legend shows the median value for the respective reservoir.

The main component of the developed image processing pipeline is the U-Net [24], a deep fully convolutional neural network used to segment growth regions of bacterial content in the assay. However, the software also utilizes classical image processing methods, such as locating the center triangle using template matching in OpenCV [25] (see Materials and Methods for details).

The developed software was evaluated on 100 CombiANT assays, having three different users taking a picture of each plate and then independently evaluating the plates using the current CombiANT software. As described in the Introduction section, this involved manually annotating CI and CP points on the plate and the triangle mark vertices, seen in Fig 1. Then, the developed image processing software was tested by processing images from all users (300 images in total), one sample shown in Fig 2. The software automatically finds the inner and outer bacteria growth zones, the white triangle-in-circle mark, and the closest distance from each triangle vertex to the inner growth zone (AB, AC, and BC). The software measured a number of distances from the circle perimeter to the outer growth zone. The median of these distances was calculated for each well and used in our analysis (A, B, and C). Software-annotated outputs from six plates from all three users are visualized in S1-S18 Figs. The results show high agreement between the manually scored assays and the users despite different lighting conditions and varying distances from the plate.

### 2.1 Outer distances

The outer distances A, B, and C, as annotated by the users, were compared to the software grading. This task was relatively more straightforward than measuring the inside distances since the latter required identifying the point in the inner growth zone closest to the corresponding triangle vertex. As shown in Figs 3-5, the software aligns with the user gradings and is more consistent having a lower standard deviation for each measurement.

**Fig 3.**
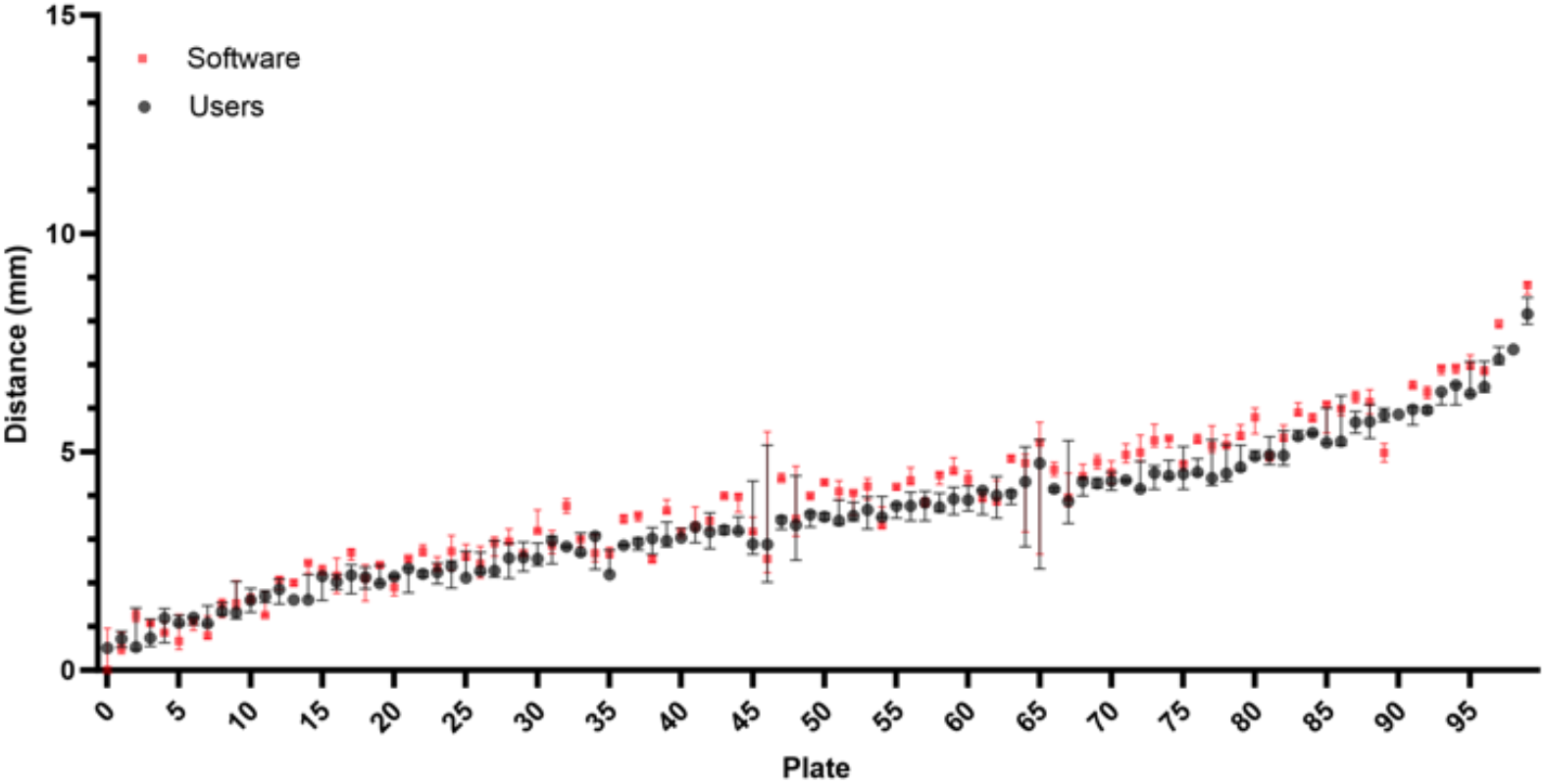
Comparison of user and software gradings for Distance A. The plates are arranged on the horizontal axis in ascending order according to the median of the measured distances (combining software and user distances). Error bars show the standard deviation of the three user and software measurements, respectively.

**Fig 4.**
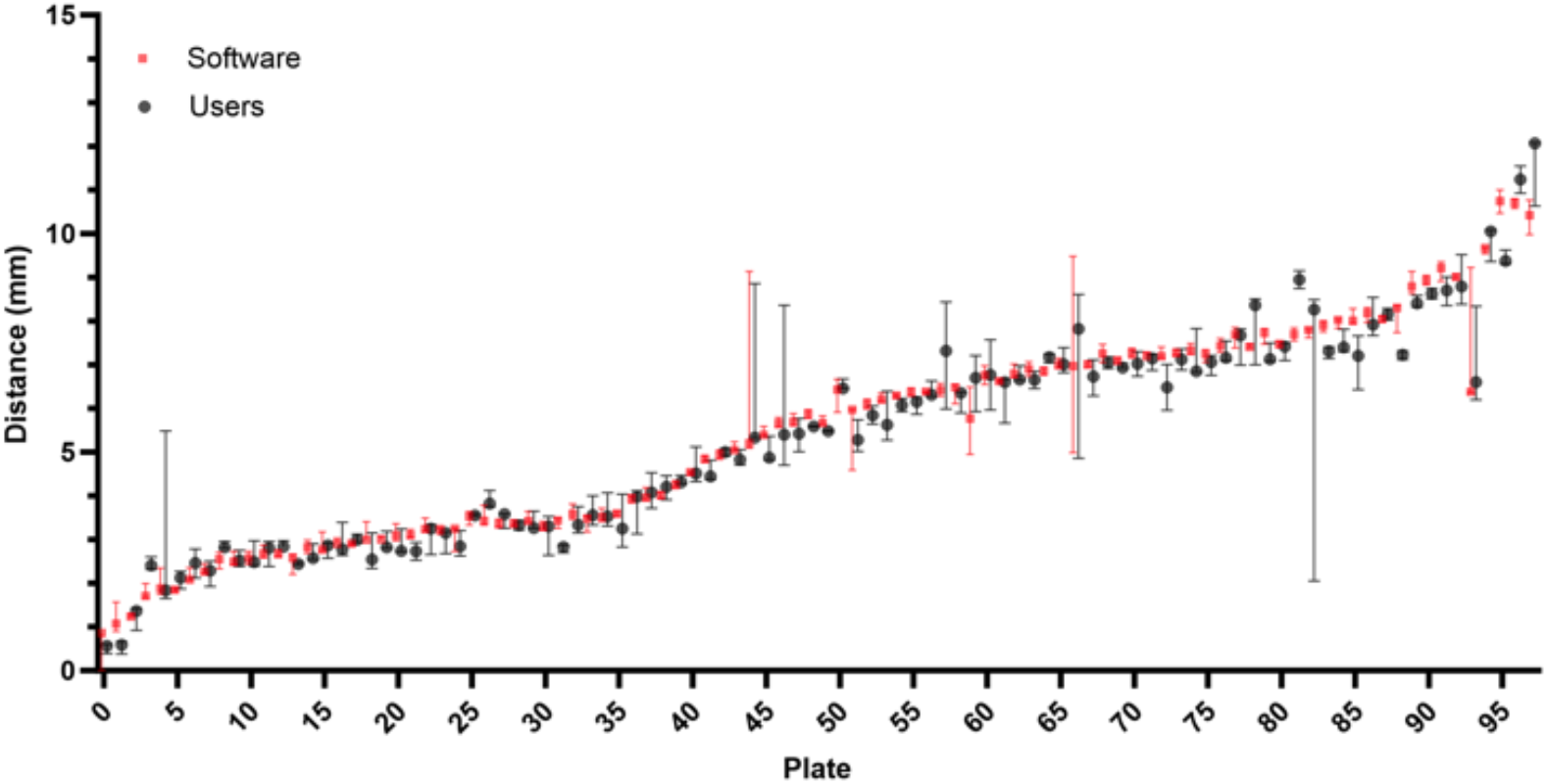
Comparison of user and software gradings for Distance B. The plates are arranged on the horizontal axis in ascending order according to the median of the measured distances (combining software and user distances). Error bars show the standard deviation of the three user and software measurements, respectively.

**Fig 5.**
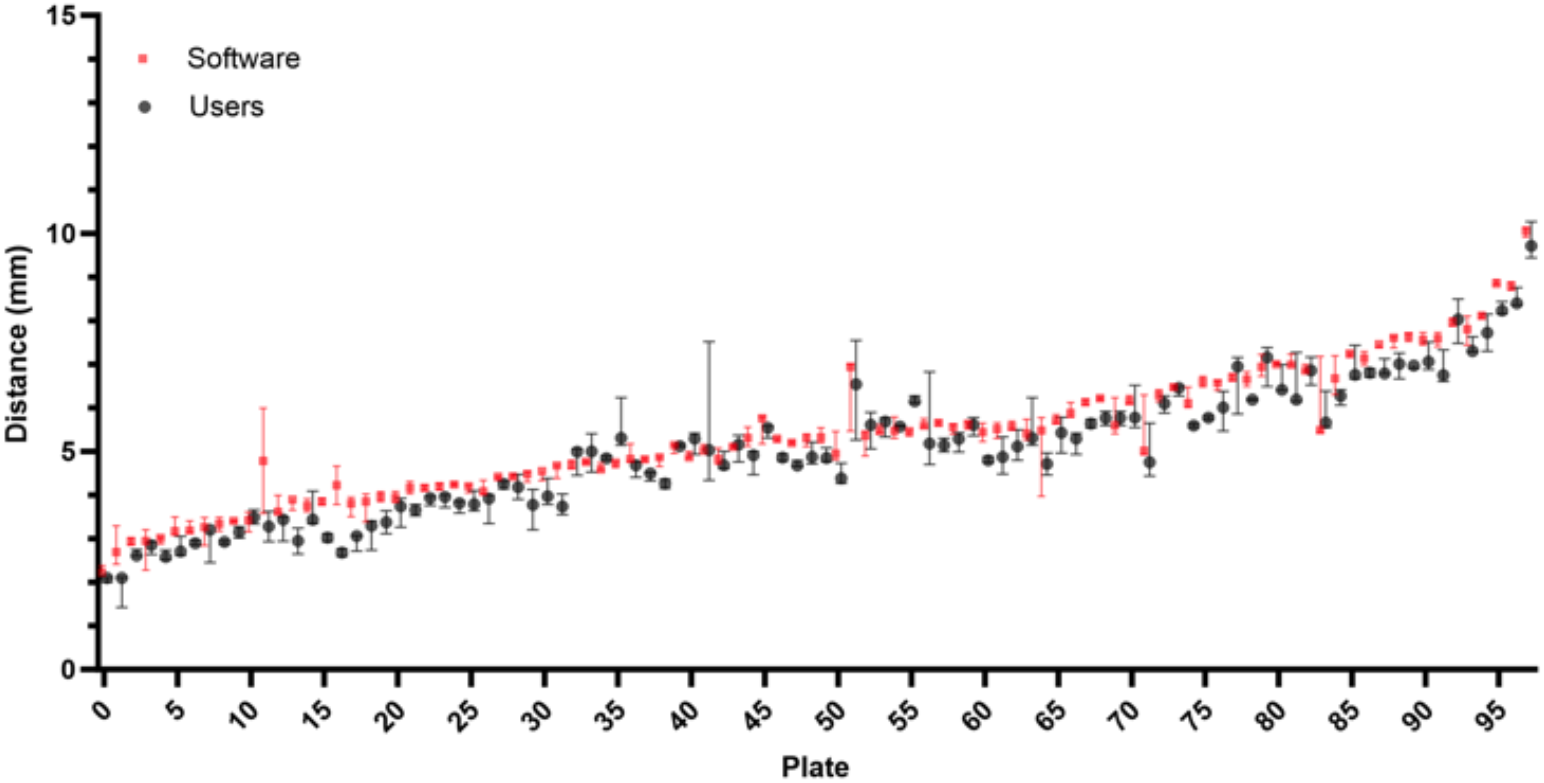
Comparison of user and software gradings for Distance C. The plates are arranged on the horizontal axis in ascending order according to the median of the measured distances (combining software and user distances). Error bars show the standard deviation of the three user and software measurements, respectively.

### 2.2 Inner distances

Estimating the inner distances (AB, AC, and BC) was more challenging for the users due to the difficulty in identifying the point closest to the triangle vertex. The three users were therefore labeled as ”Beginner,” ”Intermediate,” and ”Experienced.” The beginner user performed the first-ever grading of plates, while the experienced user had previously performed many gradings. The absolute difference between the software and user grading was calculated for each image and inner distance. As shown in Fig 6, the smallest differences were observed with the experienced user, suggesting that the developed software grading aligns more closely with the experienced user’s evaluations. The rightmost outliers correspond to discarded plates (see Discarding of plates in Materials and Methods), whose distances were still measured.

**Fig 6.**
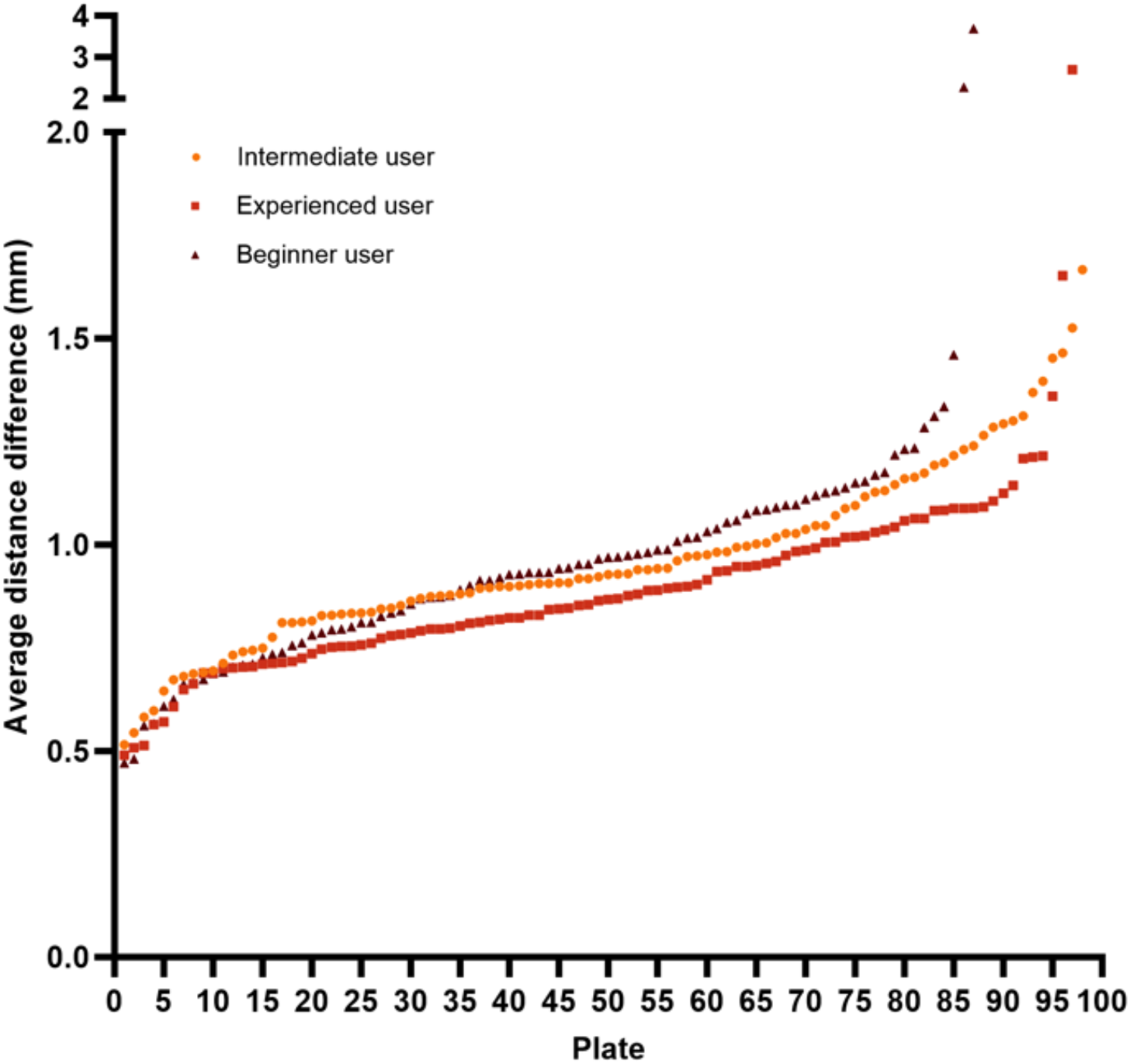
Comparison of the difference between user and software gradings for the inner distances. For each plate, the absolute difference between the user and software grading of the inner distances was calculated and then averaged over the three inner distances. The plates are arranged on the horizontal axis according to the difference between the evaluation of the experienced user and the software, in ascending order.

Furthermore, a CombiANT experiment was signaled to be discarded if any of certain criteria were met (see Discarding of plates in Materials and Methods). In the tests, the human evaluators and the software always agreed on whether to discard a plate for all test plates.

## 3 Discussion

This paper presents an image processing pipeline for automatically grading CombiAnt assays. The pipeline can process photographs from smartphone cameras, and the software can either be implemented on the client in a smartphone application, or the analysis can be performed on the server side in the cloud. The pipeline is consistent with human evaluation, robust to different imaging conditions, and fast, taking only a few seconds per plate.

The growth zone segmentation does not require any particular imaging setup or illumination, which is required by many of the classical methods [9] [10] [11]. Using augmentation during training, the model performs well even when challenged with imaging conditions not present in the trainset. If the segmentation fails, more data can easily be annotated, followed by retraining the network. In contrast, if the classical segmentation methods were used, a correction would have meant tuning existing parameters or introducing a composition of new filters and operations, which may have successfully managed to segment the failed sample but instead infer an error in another image in the trainset.

Several optimizations can be made to the software in future works. Most importantly, the template matching part could preferably be replaced with an end-to-end deep learning model that automatically finds key points, is robust to rotations shear and (if the agar plates were photographed at a tilted angle), and compensates for perspective effects. Such models could also read the marked reservoir letters on the assay (A, B, and C) and not require the CB vertex to be pointed upwards, a disadvantage of the current solution. Plenty of architectural improvements have been made since the inception of U-Net in 2014, with the development of more data- and parameter-efficient models. Most importantly, the transformer architecture [26] has also been adapted for image segmentation [27]. The model size, architecture, and subsampling step (image size where the experiment images are processed) could also be further optimized.

We hope the developed software can inspire future work developing image analysis software that processes smartphone photos of easy-to-use agar tests such as CombiANT. Such innovations will both reduce the workload and increase the robustness of the assay when assessing interactions between antibiotics, thereby facilitating work in clinical microbiology laboratories as well as in research settings.

## 4 Materials and methods

Here, the main components of the developed pipeline are described. However, for full implementation details, refer to the released software package [28], which contains documentation, a pre-trained U-Net segmentation model, annotated training data, and the test images to re-run the evaluation. The software was developed using Python 3.9.19, apart from the libraries mentioned below, the software also utilized the NumPy [29] and Pandas [30] libraries.

All images were captured using standard smartphone cameras, positioning the CombiANT assay on a flat surface and taking the photo from directly above. The raw images had a size of around 3000×3000 pixels. The image processing pipeline was written using the OpenCV [25] image processing library and Pytorch [31] and Albumentations [32] for deep neural network training. Initially, using bicubic interpolation, the software made a square center crop and downscaled the plate images to 1024×1024 pixels.

### 4.1 Segmentation of bacterial growth zones using U-Net

The pipeline employed U-Net [24], a fully convolutional artificial neural network, for segmenting the bacterial growth zones. The network was trained on segmentation masks from 1000 separate CombiANT plates not present among the plates in the evaluation. The masks were constructed by manually highlighting the bacteria boundaries using standard image processing software and then converting these images to segmentation masks. Initially, a subset of the training set was annotated, this data was then used to train a network to annotate the remaining unannotated images. The instances where the network made errors were then corrected. This strategy enabled the quick annotation of all images in the training set.

The network was trained for 1000 epochs using batch size 20, learning rate 0.0001, Cosine learning rate scheduler, and the ADAM [33] optimizer. The training time was 8 hours using the Nvidia A100 GPU. Upon receiving images from the test users to perform the evaluation, it was observed that the test images significantly differed from the training images, showing variances in background, illumination, and contrast. This caused errors in the segmentation, so the networks were retrained, adding additional augmentations, after which the model successfully segmented most of the test images. However, the contrast was manually increased using histogram equalization on a few of the evaluation images for the segmentation network to function (see Manual preprocessing).

The output segmentation mask was then inverted, and only one connected component with the largest area was filtered out, resembling the empty space without bacterial content in the center of the agar plate (the inhibition zone). The mask was then inverted again, effectively removing all holes in the segmentation mask. This process was followed by discarding connected components with a minimum area, only allowing two bacterial growth zones, ” ”outer” and ”inner”.

The segmentation was performed by first subsampling the raw images to 512×512 pixels using the ”inter area” method in OpenCV, followed by U-Net inference and thresholding, and then subsequently upsampling the output mask back to 1024×1024 pixels.

### 4.2 Alignment using template matching and grid search

Each assay was marked with a white equilateral triangle inscribed into a circle, as seen in Figs 1 and 2. This is referred to as the “triangle-in-circle”-mark or just “mark.” The corners of the triangle and the position and radius of the center circle of the mark were retrieved using a procedure utilizing template matching from the OpenCV library. First, a square center crop was extracted from the plate containing the mark, followed by applying a circular binary mask at the center, effectively zeroing out pixels outside the circle-mark perimeter. Next, the colored image was converted to a grayscale image and thresholded at the 98th percentile, obtaining a binary image revealing the bright pixels of the image, which coincided with the mark. A function was constructed, returning a binary template image of an equilateral triangle inscribed into a circle, resembling the mark, with parameters adjusting the scale (size), rotation, and line thickness. A number of these templates were generated and matched to the thresholded image utilizing an adaptive grid search outlined below.

### 4.3 Adaptive grid search

An adaptive grid search was used to obtain the optimal parameters of the template that best match the thresholded image containing the mark. The procedure first performed a search using scale, then rotation. The reason for starting with scale is that the correct scale, the circle part of the mark, overlaps with the template regardless of orientation. The procedure searched for a number of values above and below a start value with a predetermined step size (0.01 and 1 for rotation and scale, respectively), obtaining the best fit, location, and value (scale or rotation). The fit and location were obtained from the OpenCV template matching algorithm. The grid search algorithm used is outlined in Algorithm 1.

#### Algorithm 1

Adaptive Grid Search for Optimal Template Matching Parameters

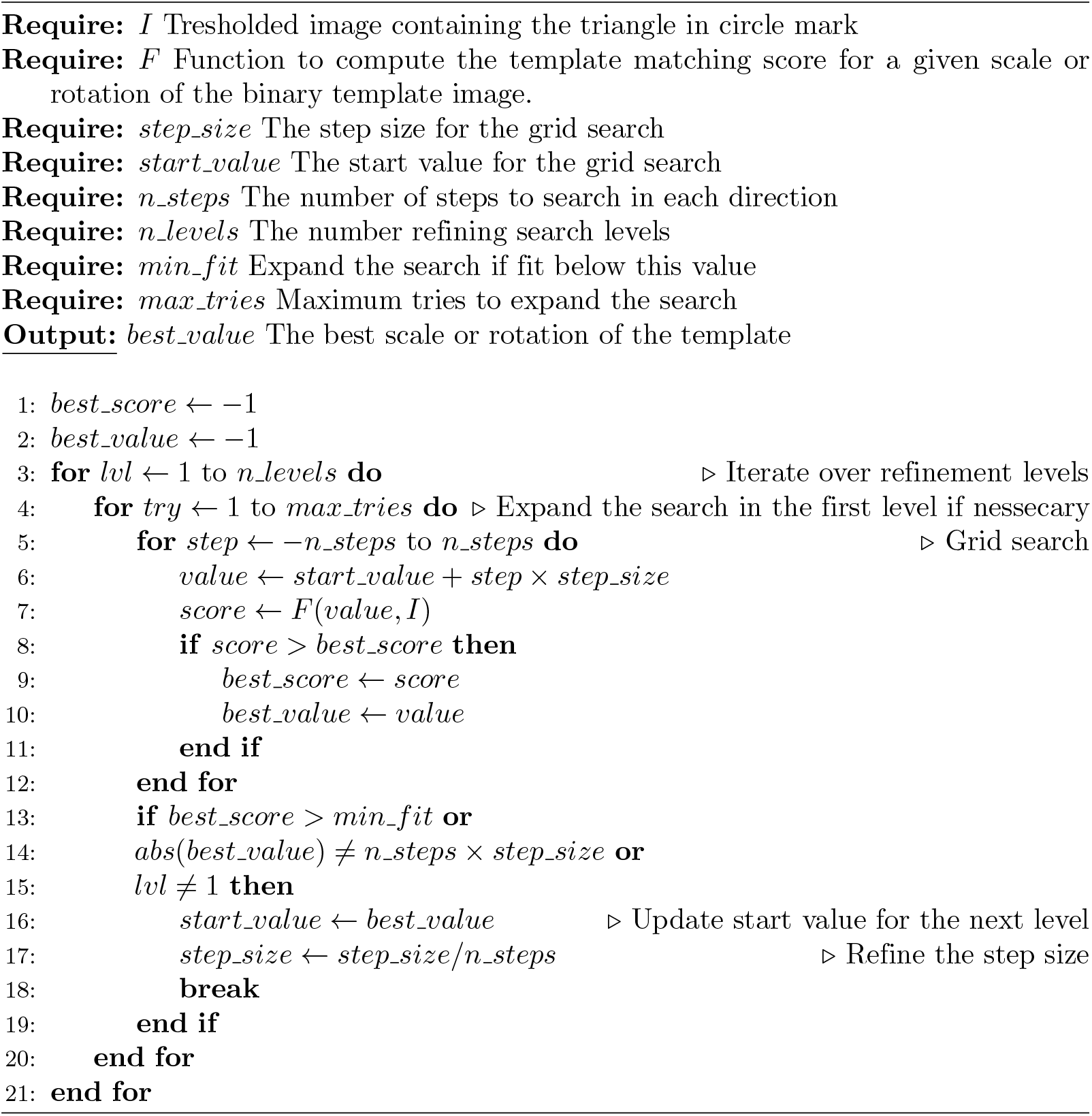

#### 4.3.1 Expanding search

If the value with the best fit was the minimum or maximum value in the grid search, the search was retried with the number of search values doubled, using the identical step size. The search was also expanded if the best fit was below a predetermined threshold.

#### 4.3.2 Refining Search

Once the approximate best-fit value was identified, a finer search was performed around that optimal value with a smaller step size. This refining search was executed once by default but could be expanded with more levels. The step size was adjusted to search around the best-fit value, extending from the previous value to the next value (relative to the best-fit on the previous level).

Following this grid search, the triangle-in-circle mark was outlined on each test sample with a black border, as seen in Fig 2 and S1-S18 Figs.

### 4.4 Measurement of distances

After the triangle vertices and position of the circle-mark perimeter were found using the template matching step, distances were measured on the assay. First, the binary mask from the U-Net processing, where connected components resembled bacteria content, was converted to contours using the OpenCV ”drawContours”-function. The measurements were performed by drawing straight lines on the binary mask containing the contours from key points of interest until the line encountered a contour. The end point of the line, and thus the distances, was obtained by finding the point where the contour and line intersected.

#### 4.4.1 Inner growth zone

The minimum distance along a straight line from each triangle vertex to the inner growth zone was measured and visualized as green lines in Fig 2 and S1-S18 Figs. The distances are shown as AB, AC and BC in the legend, corresponding to the respective triangle vertex. The measurement was obtained by drawing a number of test lines from the corresponding triangle vertex to the inner growth zone. The line with the shortest distance was retained as the minimum distance.

#### 4.4.2 Outer growth zone

A total of 55 distances were measured per well, drawing lines perpendicular to the circle-mark perimeter to the rim of the outer growth zone. A 20-degree padding was added on each side of every triangle-mark vertex since the combination concentration is larger there (two antibiotic wells intersect). The outer distances are supposed to measure susceptibility to one antibiotic only. The median of the distances per well was calculated and shown as A, B and, C in the figure legend in Fig 2 and S1-S18 Figs.

#### 4.4.3 Millimeter conversion

All distance measurements were initially obtained in pixels. Then, the resolution *R*(*i*) was calculated for image *i* ∈ *test_images* using the radius of the circle-mark known from the fabrication (*r*_*mm*_ = 20.5*mm*), and the radius of the current image *r*(*i*)_*px*_ in pixels, obtained from the template matching step.

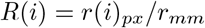

The resolution for the images was at 10.069 +-1.604 px/mm (mean, standard deviation). Using this information, all pixel distances *d*_*px*_ were converted to millimeters *d*_*mm*_ for all images *i, d*_*mm*_ = *d*_*px*_*/R*(*i*). If the plates were photographed from a far distance, there could be a precision issue, but this was not a problem in practice.

### 4.5 Manual preprocessing

One of the users (User 3) did not align the plates before photographing, pointing the CB vertex up. We manually cropped and rotated these images. U-net segmentation failed on five images from User 1 and 16 images from User 3. We manually increased the contrast of these images, converting to YCrCb color space, and increased the contrast in the Y-channel (which corresponds to the brightness) using histogram equalization.

### 4.6 Discarding of plates

A plate was discarded when any of three criteria were met:

- The inner growth zone grew outside of the triangle-mark interaction zone.
- The outer growth zone crossed the circle mark perimeter at a point where outer distances were measured (not considering points within the 20-degree padding from each triangle vertex).
- There was no center growth zone, it has disappeared due to the high synergetic effect of the antibiotics.

These conditions were detected in the pipeline with added discard warnings, shown as ”inner”, ”outer”, and ”no_island” on the legend. These plates were still processed and distances measured, but with the warning attached, see S18-S20 Figs.

### 4.7 Inference time

Processing one sample took around 7 seconds, with U-net inference taking 0.0376+-0.0283 seconds (1.039+-0.446. seconds when running on the CPU) and template matching and grid search taking 7.240+-1.660 seconds.

### 4.8 Use of artificial intelligence tools and technologies

The tools Grammarly and ChatGPT were used for grammar checking, as a synonym book, and as rephrasing tools when writing this article. No original content was generated by the models. Furthermore, the coding assistant GitHub Copilot was used to generate type annotations and docstrings for the software in the released replication package.

## Data Availability

The authors confirm that all data underlying the findings are fully available without restriction. A replication package is available at https://doi.org/10.5281/zenodo.13893289 containing all image data and software to reproduce the experiments, generate output metrics and build the graphs in the article.

## 5 Supporting information

### 5.1 Output images

Six plates from the study are shown below, photographed by each of the three users and annotated with an overlay created by our CombiANT reader. Our developed software finds the circle mark with the inscribed triangle using template matching, outlined with a black border, and the edges of the bacterial growth zones, outlined in red. The software measures 55 distances from each reservoir to the outer growth zone (blue lines) and automatically finds the closest distance to the inner growth zone from every triangle vertex (green lines). The legend shows the calculated distances in millimeters with two decimals. For the outer distances, the legend shows the median value for the respective reservoir.

**S1 Fig.**
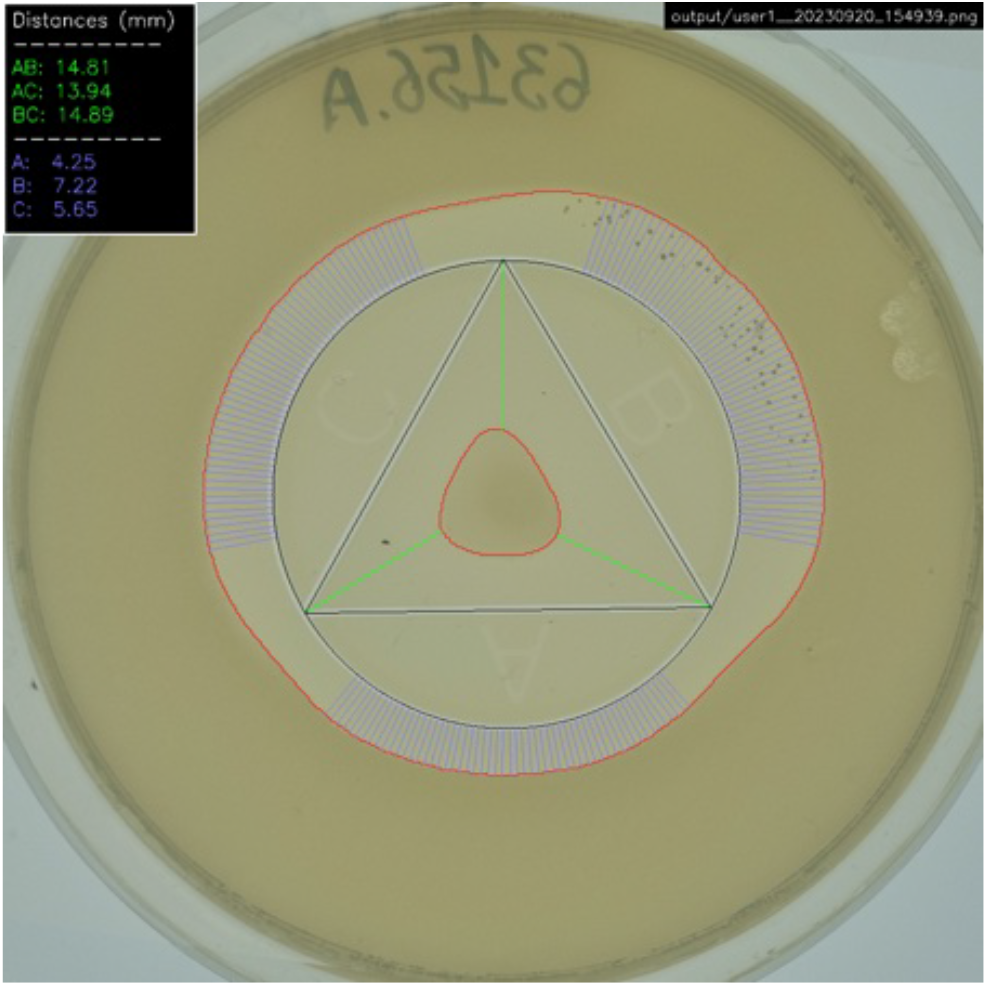
Plate 1 photographed by User 1.

**S2 Fig.**
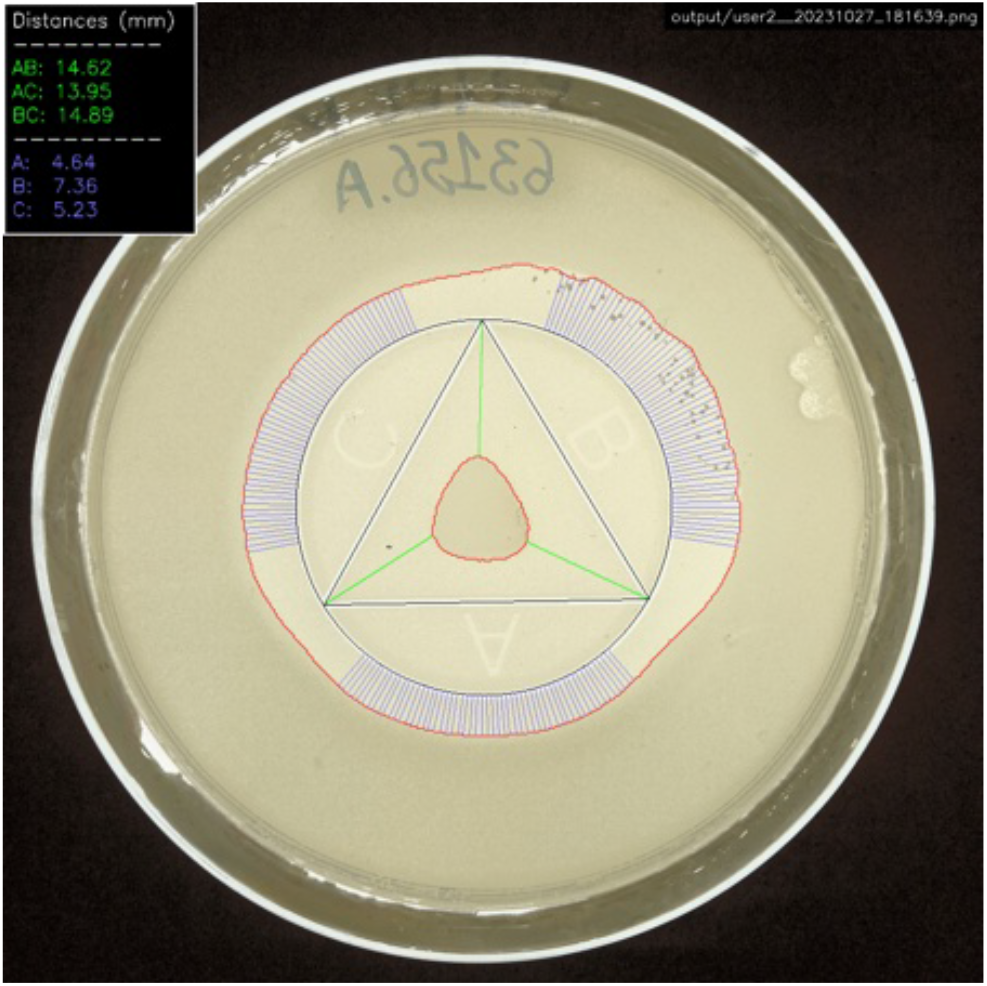
Plate 1 photographed by User 2.

**S3 Fig.**
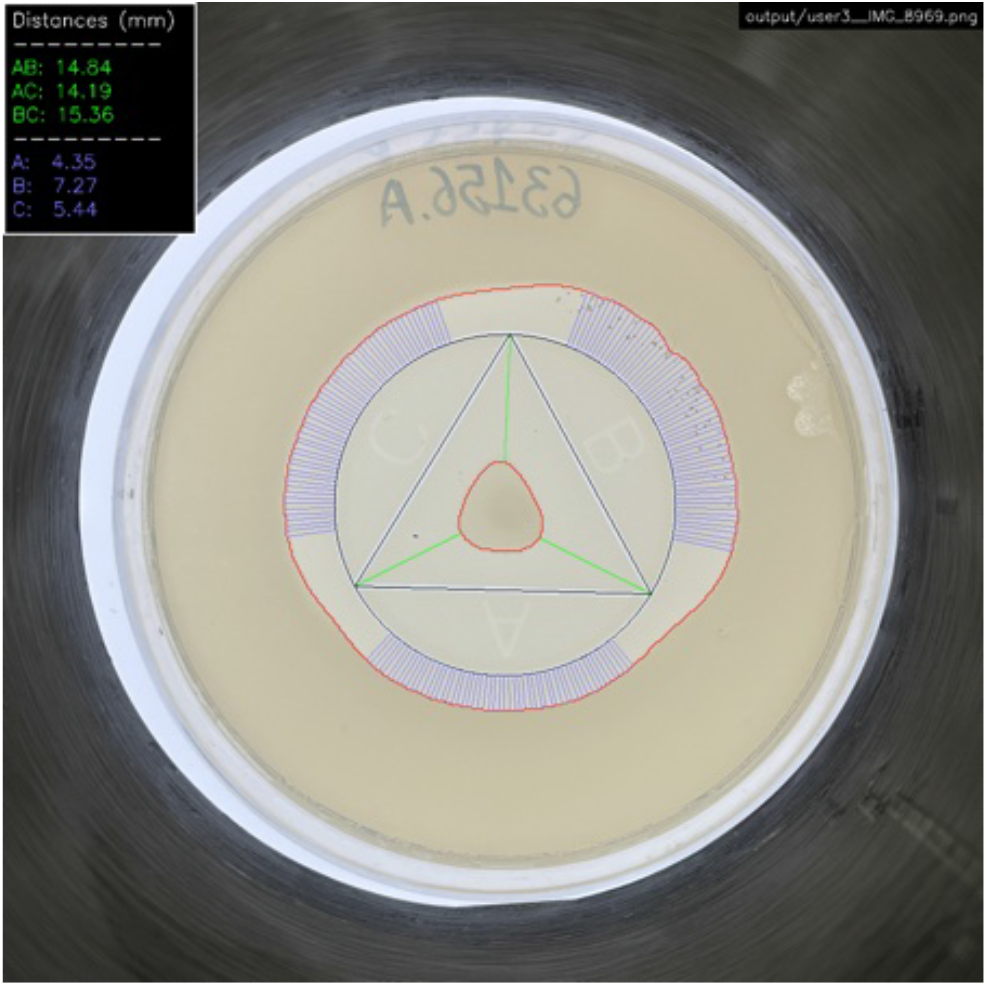
Plate 1 photographed by User 3.

**S4 Fig.**
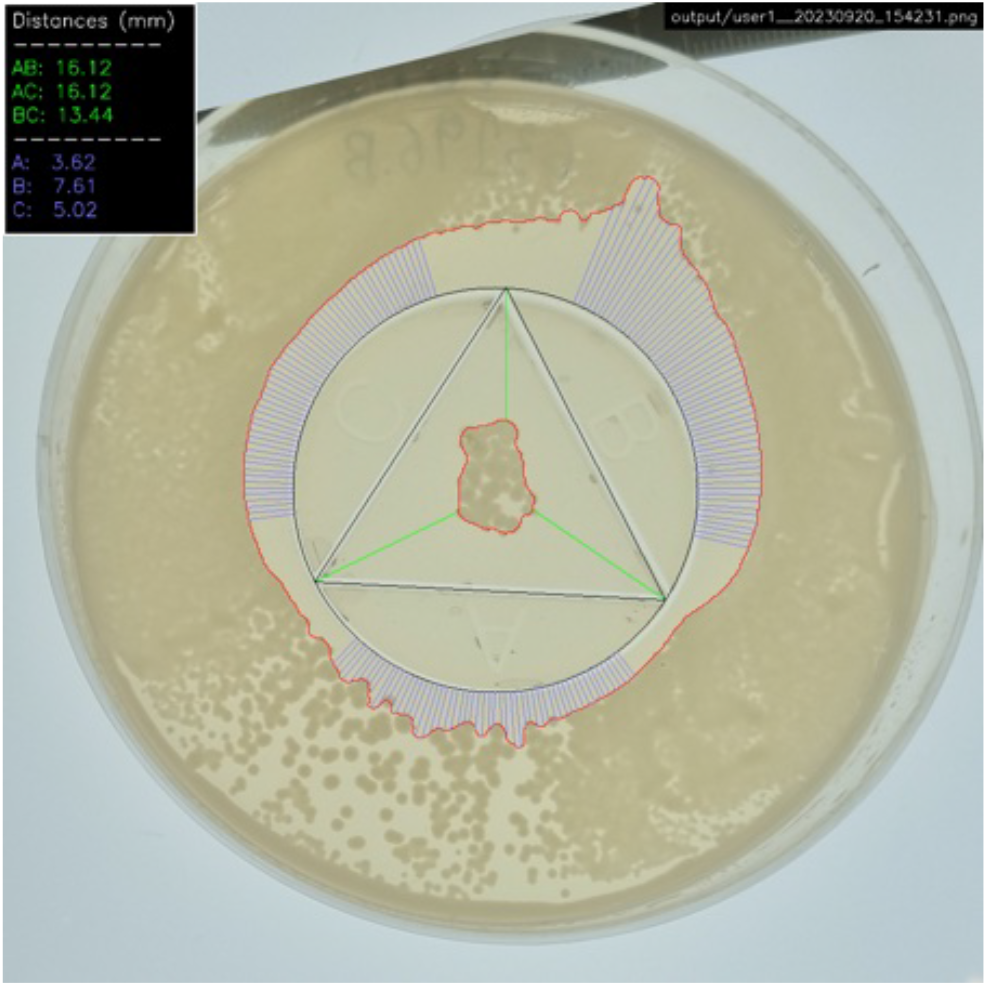
Plate 2 photographed by User 1.

**S5 Fig.**
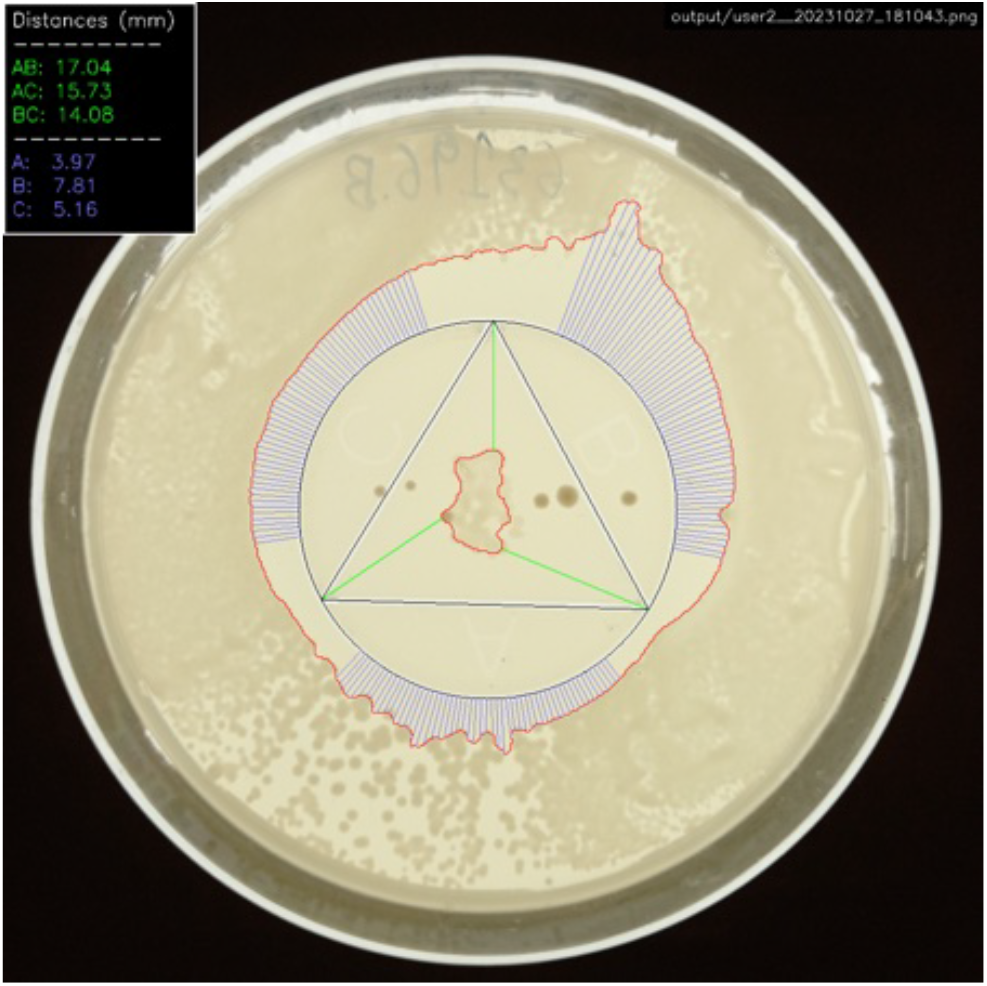
Plate 2 photographed by User 2.

**S6 Fig.**
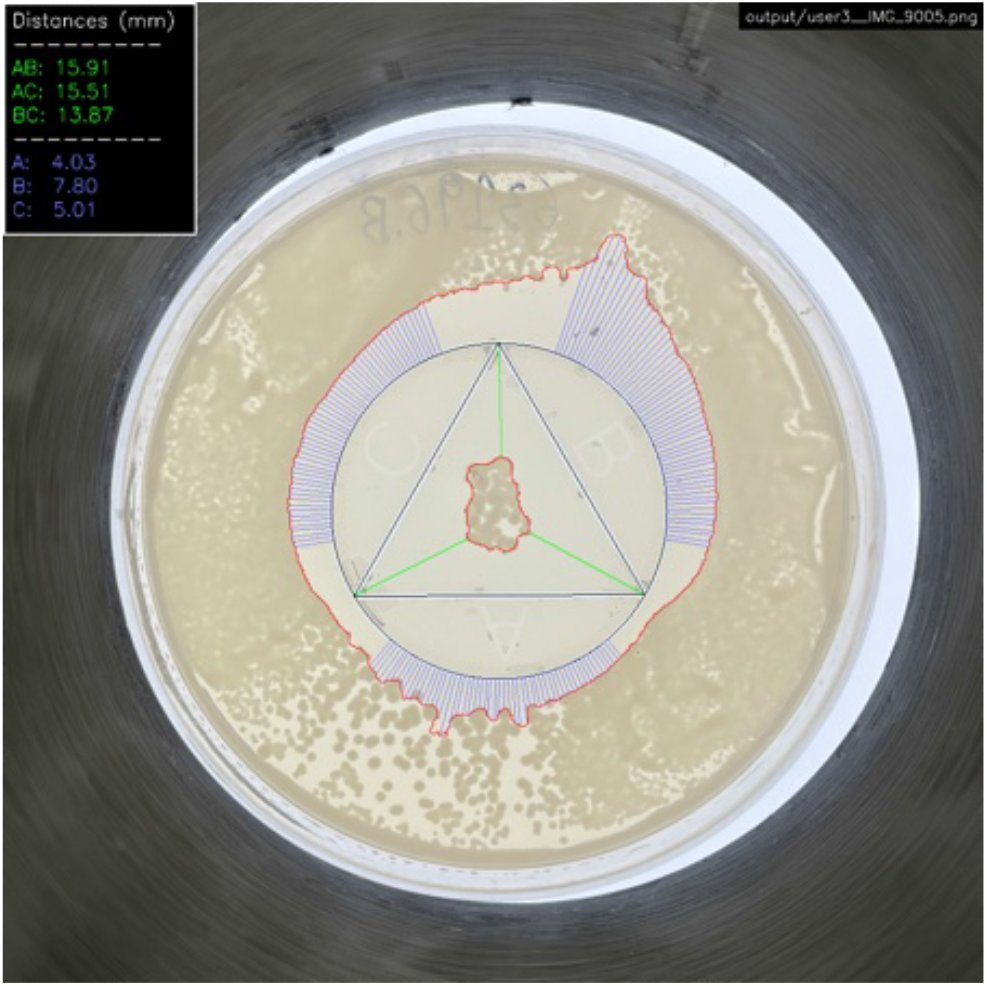
Plate 2 photographed by User 3.

**S7 Fig.**
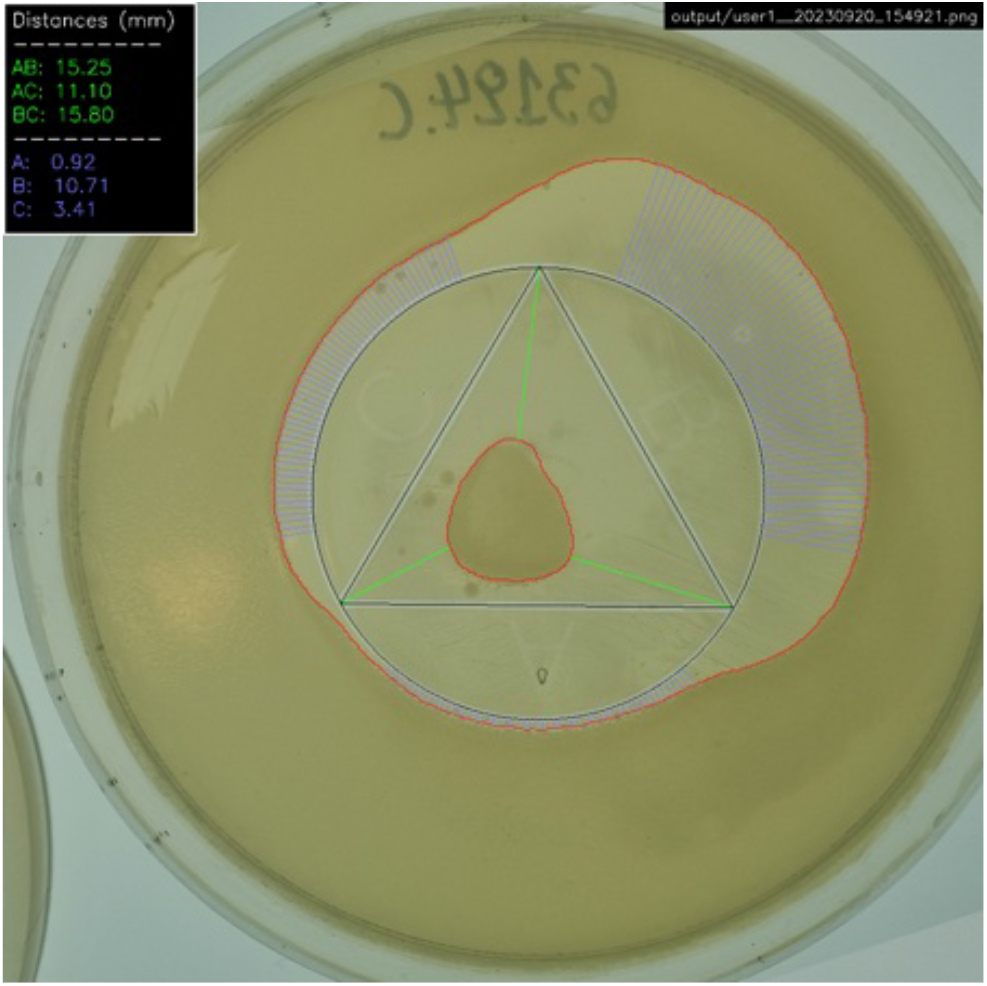
Plate 3 photographed by User 1.

**S8 Fig.**
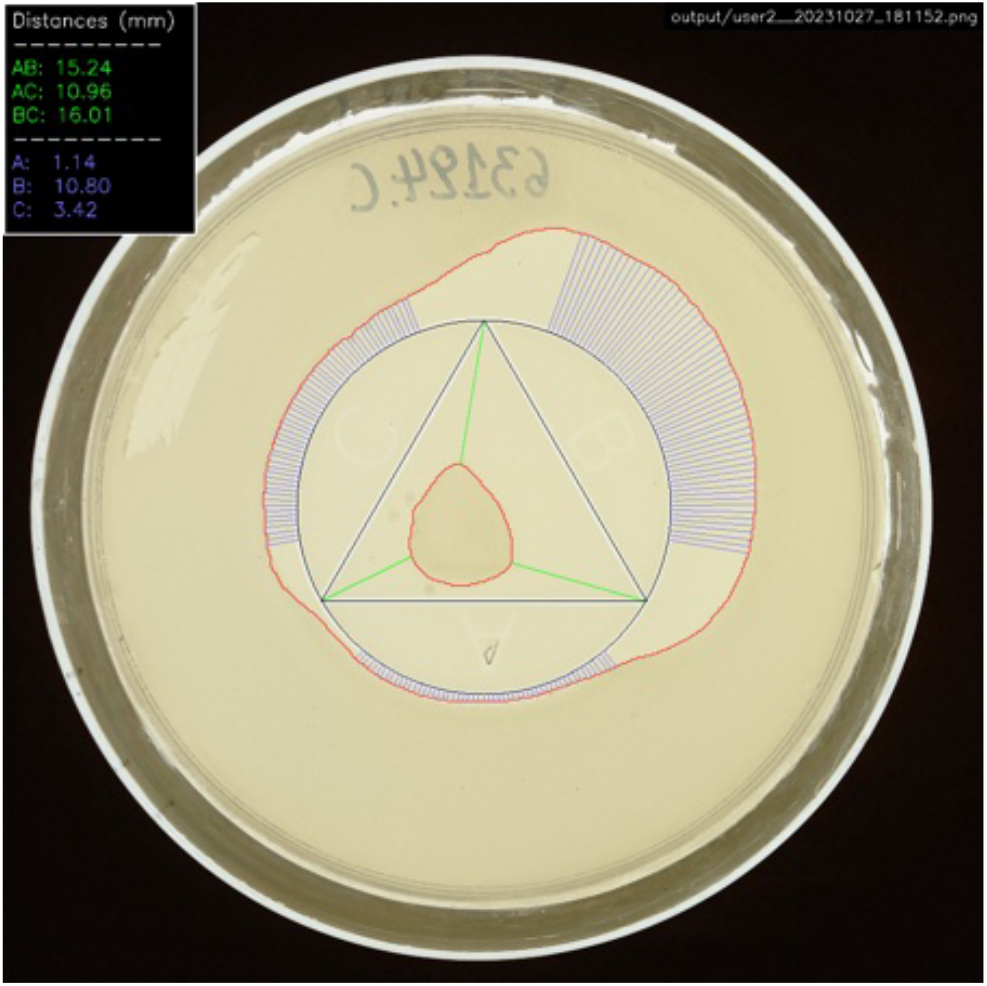
Plate 3 photographed by User 2.

**S9 Fig.**
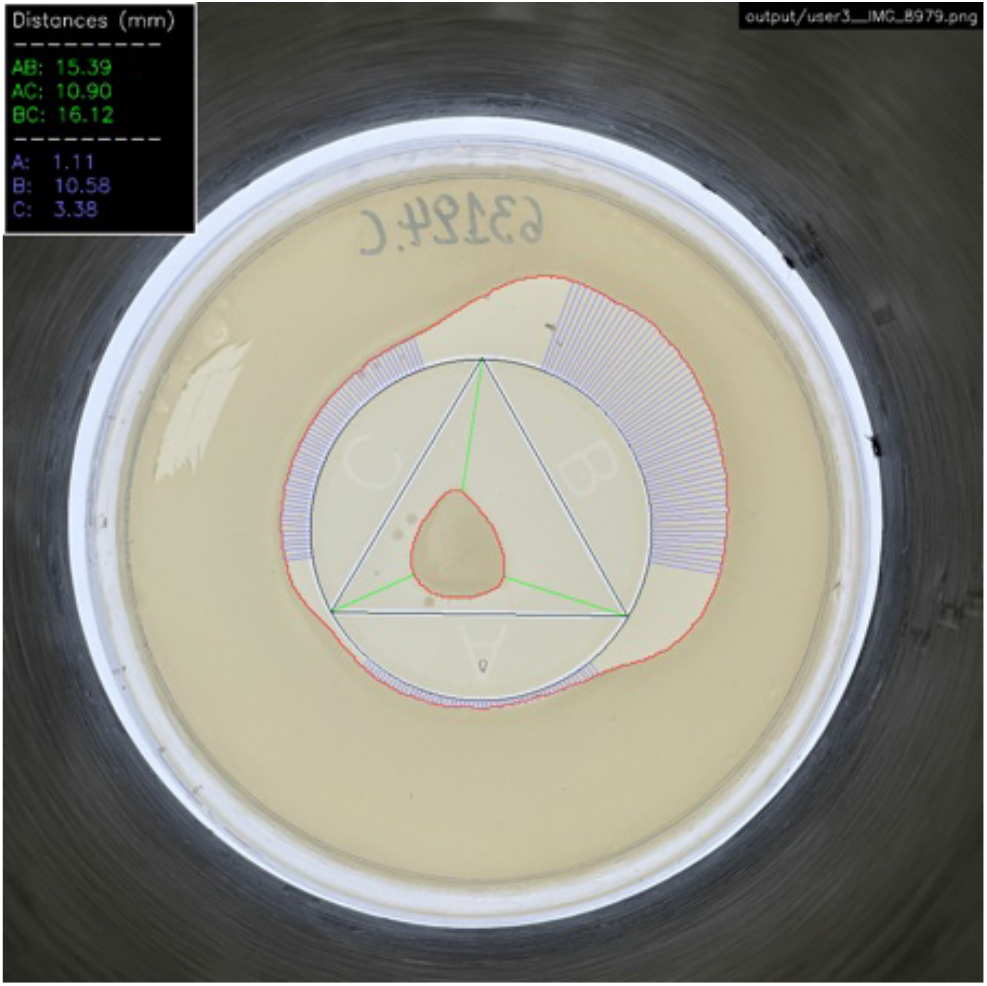
Plate 3 photographed by User 3.

**S10 Fig.**
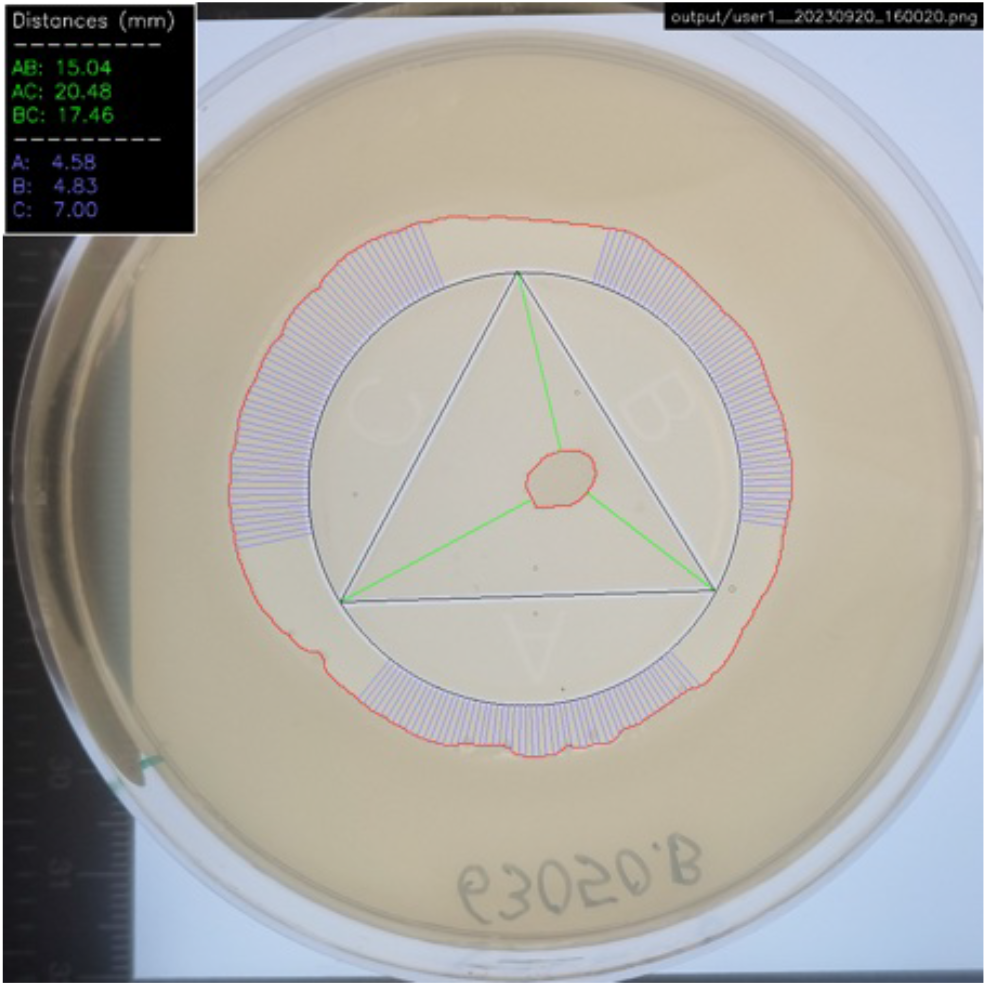
Plate 4 photographed by User 1.

**S11 Fig.**
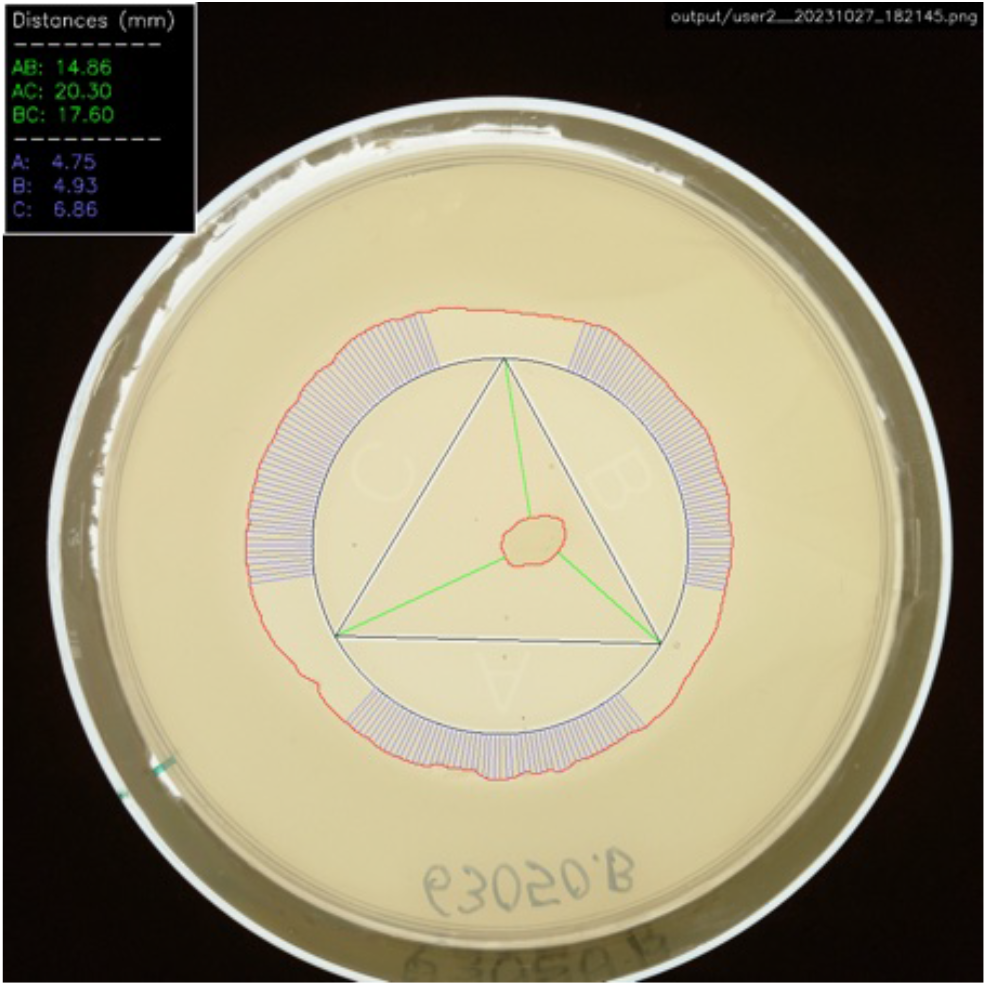
Plate 4 photographed by User 2.

**S12 Fig.**
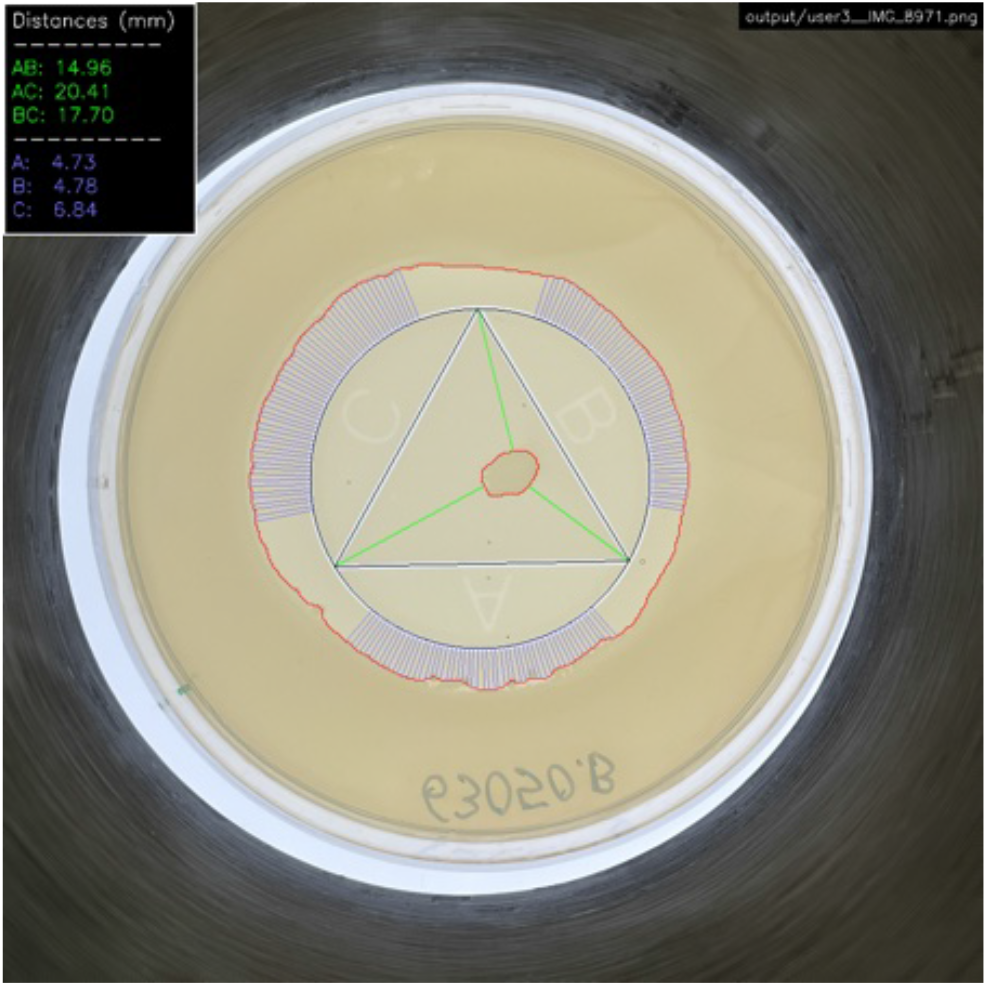
Plate 4 photographed by User 3.

**S13 Fig.**
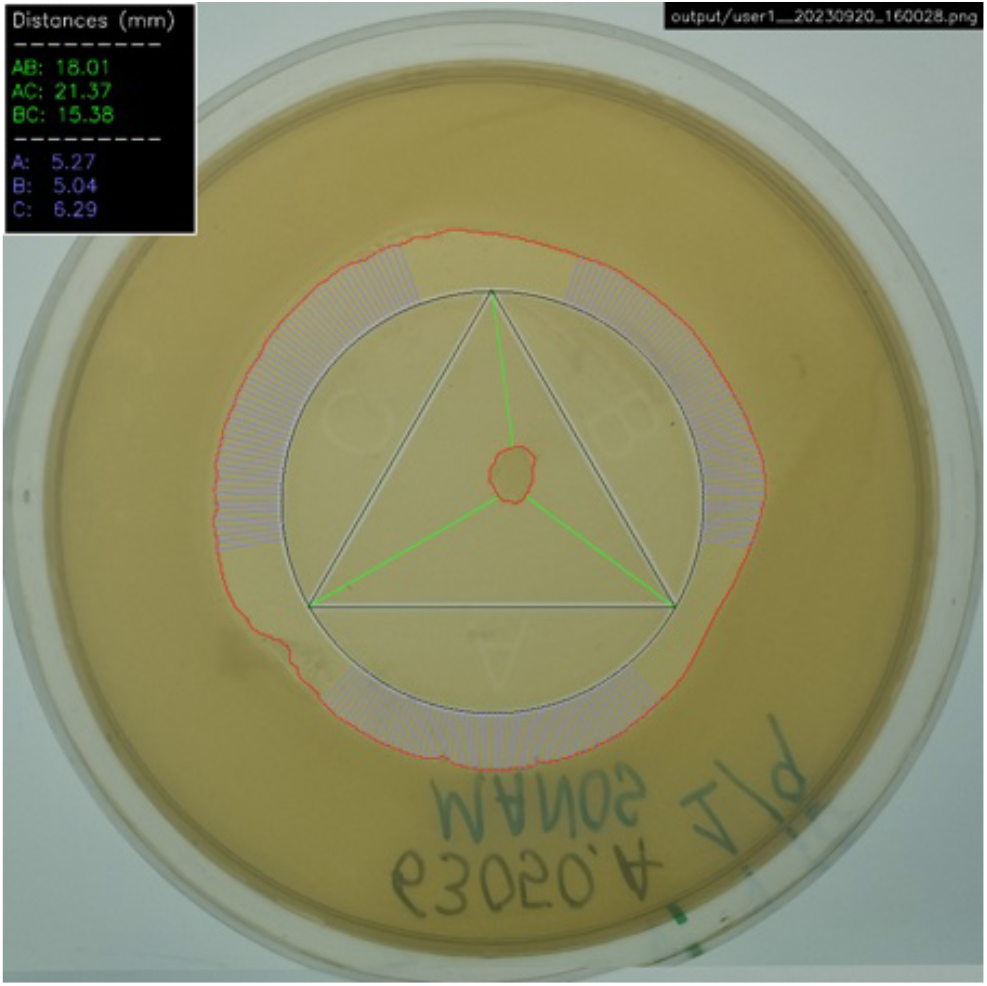
Plate 5 photographed by User 1.

**S14 Fig.**
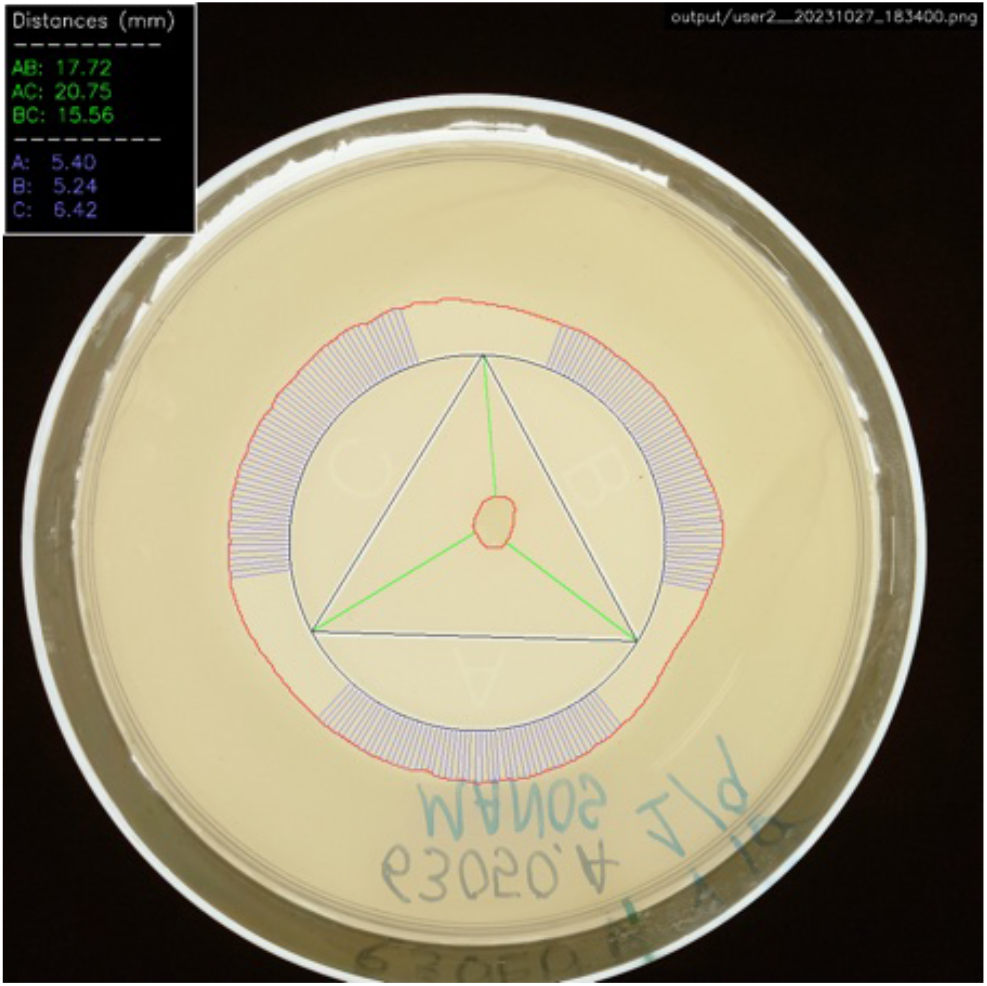
Plate 5 photographed by User 2.

**S15 Fig.**
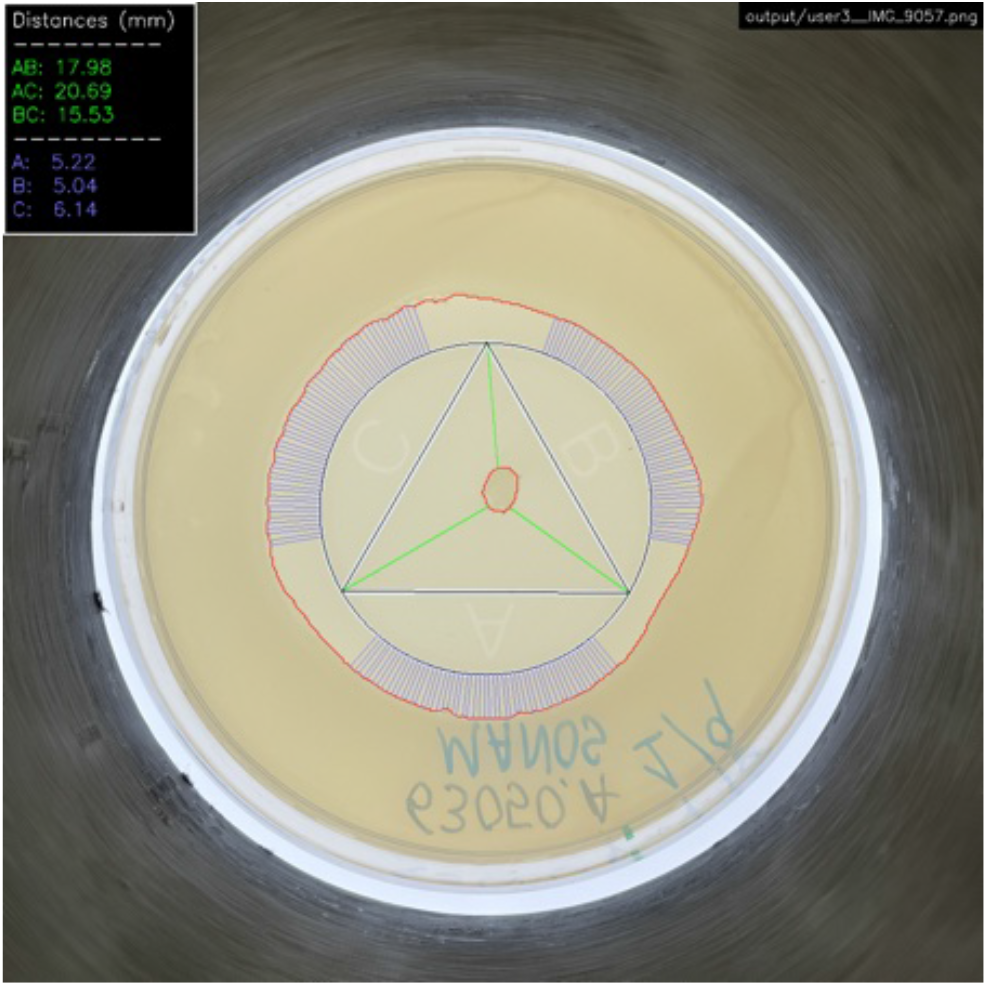
Plate 5 photographed by User 3.

**S16 Fig.**
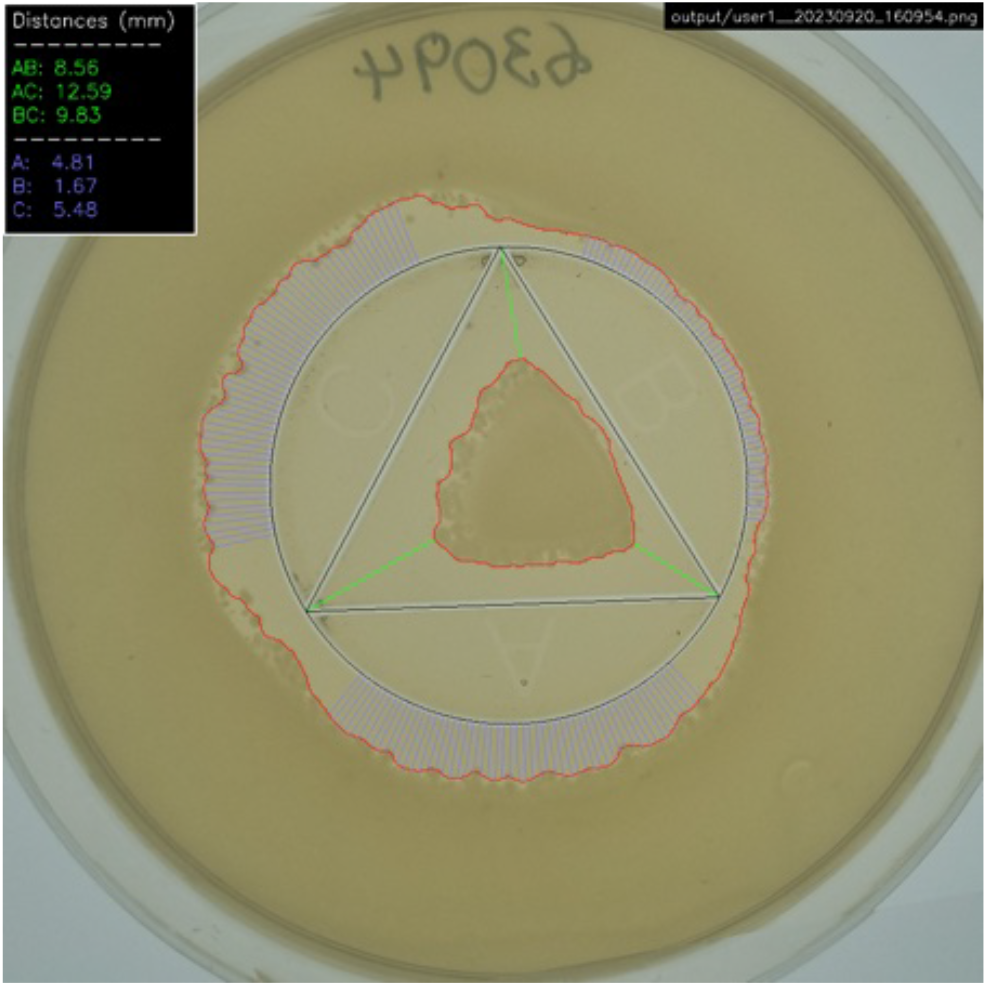
Plate 6 photographed by User 1.

**S17 Fig.**
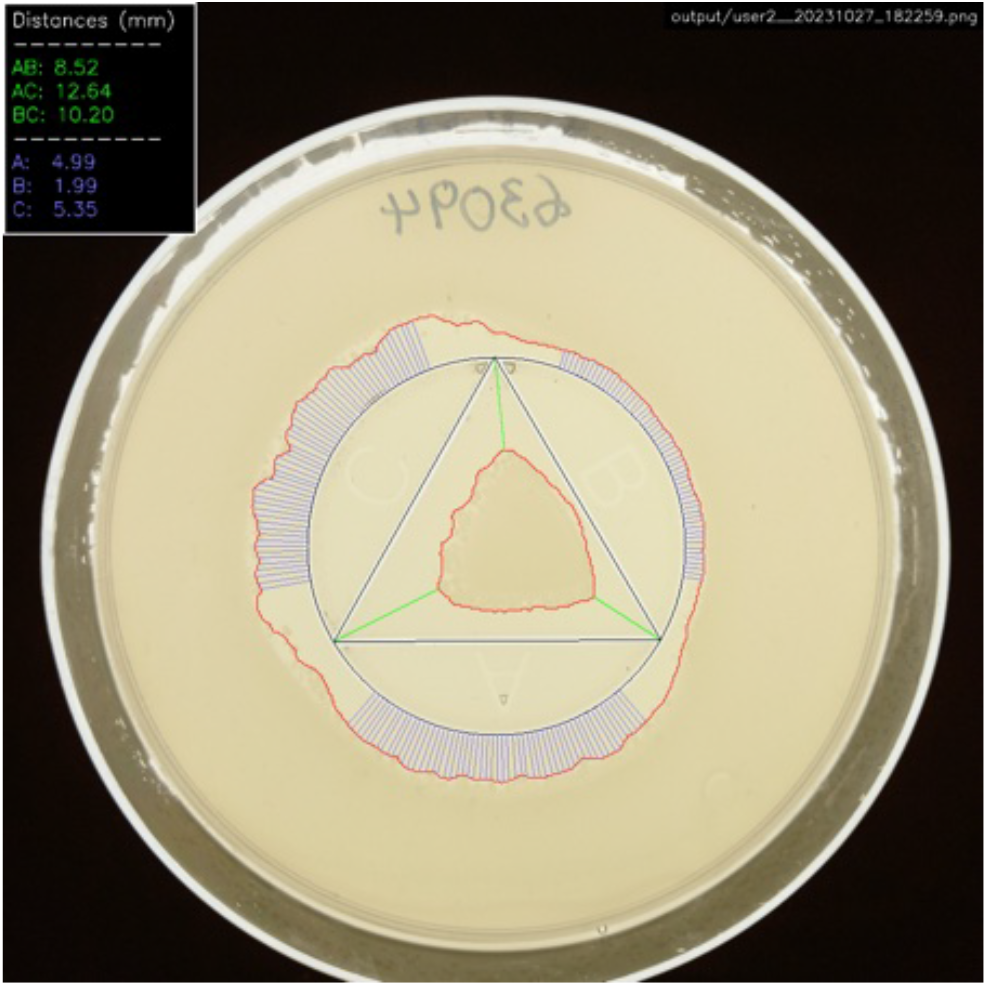
Plate 6 photographed by User 2.

**S18 Fig.**
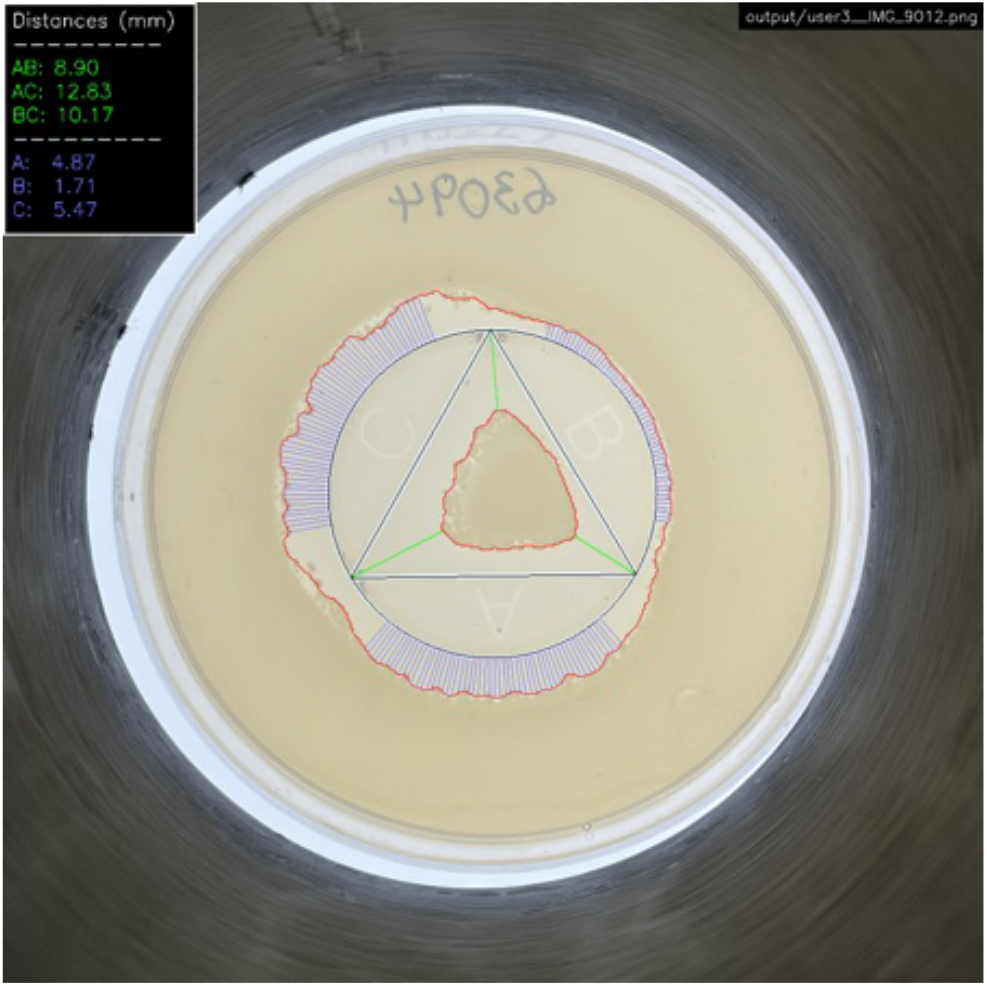
Plate 6 photographed by User 3.

### 5.2 Examples of discarded plates

**S19 Fig.**
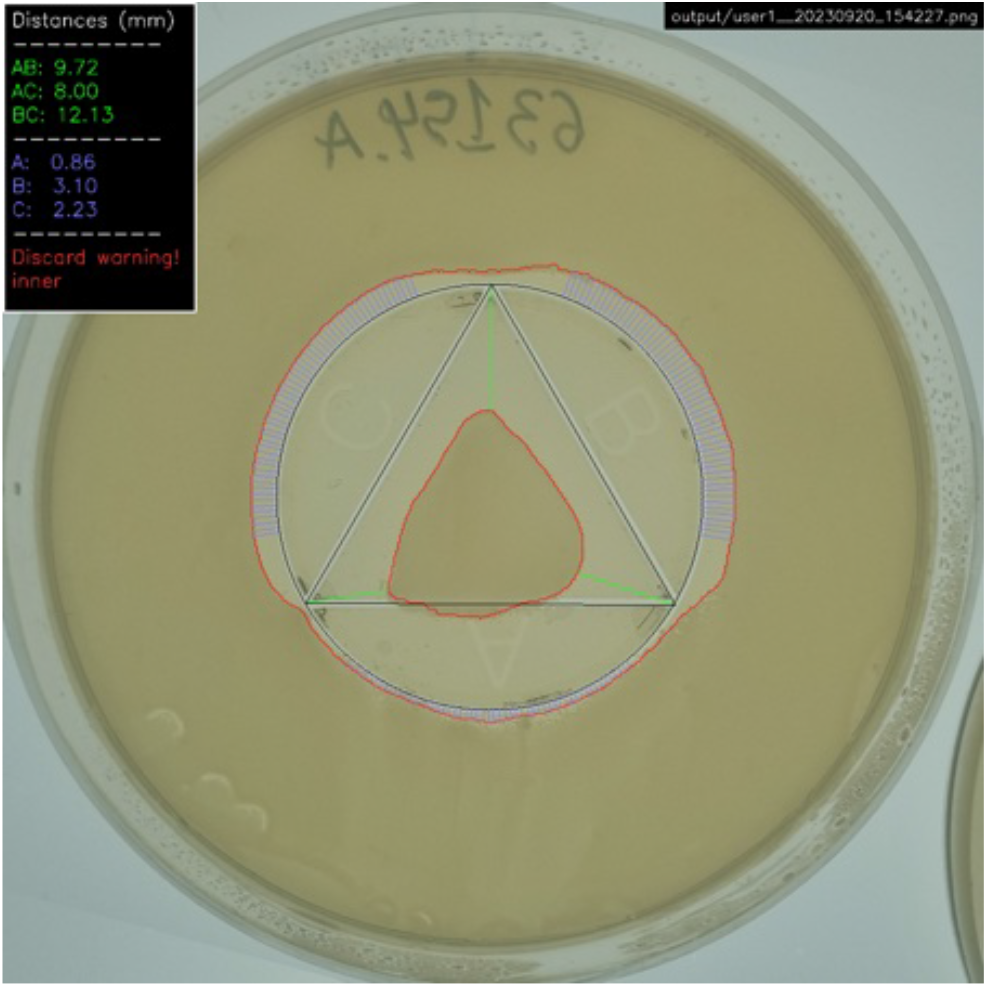
Inner discard. Discarded plate due to inner growth zone intersecting with the triangle-mark.

**S20 Fig.**
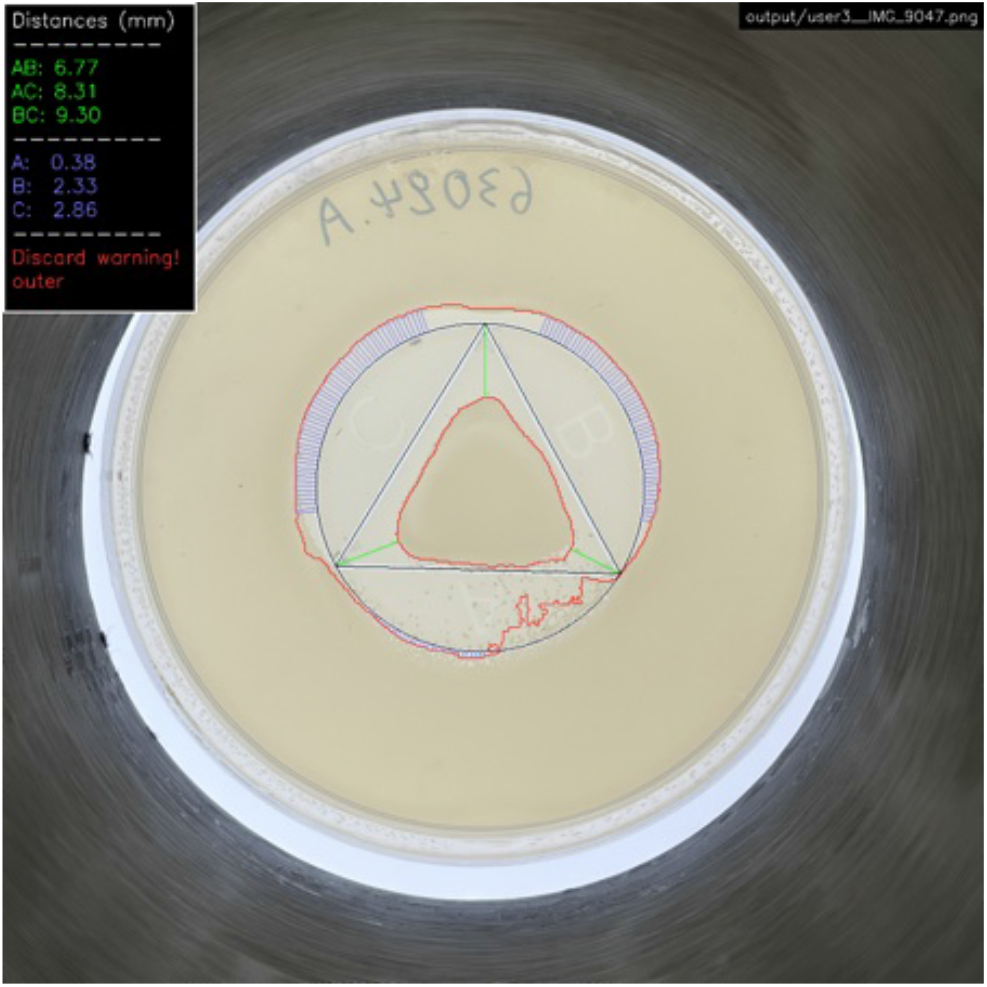
Outer discard. Discarded plate due to the outer growth zone growing past the circle mark.

## References

1. Murray CJL, Ikuta KS, Sharara F, Swetschinski L, Robles Aguilar G, Gray A, et al. Global burden of bacterial antimicrobial resistance in 2019: a systematic analysis. The Lancet. 2022;399(10325):629–655. doi:10.1016/S0140-6736(21)02724-0.

2. Tang PC, Sánchez-Hevia DL, Westhoff S, Fatsis-Kavalopoulos N, Andersson DI. Within-species variability of antibiotic interactions in Gram-negative bacteria. mBio. 2024;15(3):e00196–24. doi:10.1128/mbio.00196-24.

3. Roemhild R, Bollenbach T, Andersson DI. The physiology and genetics of bacterial responses to antibiotic combinations. Nature Reviews Microbiology. 2022;20(8):478–490.

4. Fatsis-Kavalopoulos N, Roelofs L, Andersson DI. Potential risks of treating bacterial infections with a combination of, B-lactam and aminoglycoside antibiotics: A systematic quantification of antibiotic interactions in E. coli blood stream infection isolates. EBioMedicine. 2022;78.

5. Cacace E, Kim V, Varik V, Knopp M, Tietgen M, Brauer-Nikonow A, et al. Systematic analysis of drug combinations against Gram-positive bacteria. Nature Microbiology. 2023;8(11):2196–2212.

6. Brochado AR, Telzerow A, Bobonis J, Banzhaf M, Mateus A, Selkrig J, et al. Species-specific activity of antibacterial drug combinations. Nature. 2018;559(7713):259–263.

7. Fatsis-Kavalopoulos N, Roemhild R, Tang PC, Kreuger J, Andersson DI. CombiANT: Antibiotic interaction testing made easy. PLoS biology. 2020;18(9):e3000856.

8. Zhang J, Li C, Rahaman MM, Yao Y, Ma P, Zhang J, et al. A comprehensive review of image analysis methods for microorganism counting: from classical image processing to deep learning approaches. Artificial Intelligence Review. 2022; p. 1–70.

9. Arora P, Tewary S, Krishnamurthi S, Kumari N. An experimental setup and segmentation method for CFU counting on agar plate for the assessment of drinking water. Journal of Microbiological Methods. 2023;214:106829.

10. Brugger SD, Baumberger C, Jost M, Jenni W, Brugger U, Mühlemann K. Automated counting of bacterial colony forming units on agar plates. PloS one. 2012;7(3):e33695.

11. Hogekamp L, Hogekamp SH, Stahl MR. Experimental setup and image processing method for automatic enumeration of bacterial colonies on agar plates. PLoS One. 2020;15(6):e0232869.

12. Chen WB, Zhang C. An automated bacterial colony counting and classification system. Information Systems Frontiers. 2009;11:349–368.

13. Chiang PJ, Tseng MJ, He ZS, Li CH. Automated counting of bacterial colonies by image analysis. Journal of microbiological methods. 2015;108:74–82.

14. Boukouvalas DT, Prates RA, Leal CRL, de Araújo SA. Automatic segmentation method for CFU counting in single plate-serial dilution. Chemometrics and Intelligent Laboratory Systems. 2019;195:103889.

15. Smith P, Schuster M. Inexpensive apparatus for high-quality imaging of microbial growth on agar plates. Frontiers in Microbiology. 2021;12:689476.

16. Andreini P, Bonechi S, Bianchini M, Mecocci A, Scarselli F. A deep learning approach to bacterial colony segmentation. In: Artificial Neural Networks and Machine Learning–ICANN 2018: 27th International Conference on Artificial Neural Networks, Rhodes, Greece, October 4-7, 2018, Proceedings, Part III 27. Springer; 2018. p. 522–533.

17. Ferrari A, Lombardi S, Signoroni A. Bacterial colony counting with convolutional neural networks in digital microbiology imaging. Pattern Recognition. 2017;61:629–640.

18. Shamash M, Maurice CF. OnePetri: accelerating common bacteriophage Petri dish assays with computer vision. Phage. 2021;2(4):224–231.

19. Nagy SÁ, Makrai L, Csabai I, Tőzsér D, Szita G, Solymosi N. Bacterial colony size growth estimation by deep learning. BMC microbiology. 2023;23(1):307.

20. Pascucci M, Royer G, Adamek J, Asmar MA, Aristizabal D, Blanche L, et al. AI-based mobile application to fight antibiotic resistance. Nature communications. 2021;12(1):1173.

21. Alonso CA, Domínguez C, Heras J, Mata E, Pascual V, Torres C, et al. Antibiogramj: A tool for analysing images from disk diffusion tests. Computer methods and programs in biomedicine. 2017;143:159–169.

22. Fough F, Janjua G, Zhao Y, Don AW. Predicting and Identifying Antimicrobial Resistance in the Marine Environment Using AI & Machine Learning Algorithms. In: 2023 IEEE International Workshop on Metrology for the Sea; Learning to Measure Sea Health Parameters (MetroSea). IEEE; 2023. p. 121–126.

23. LeCun Y, Bengio Y, Hinton G. Deep learning. nature. 2015;521(7553):436–444.

24. Ronneberger O, Fischer P, Brox T. U-net: Convolutional networks for biomedical image segmentation. In: Medical image computing and computer-assisted intervention–MICCAI 2015: 18th international conference, Munich, Germany, October 5-9, 2015, proceedings, part III 18. Springer; 2015. p. 234–241.

25. Bradski G. The OpenCV Library. Dr Dobb’s Journal of Software Tools. 2000;.

26. Dosovitskiy A, Beyer L, Kolesnikov A, Weissenborn D, Zhai X, Unterthiner T, et al. An Image is Worth 16×16 Words: Transformers for Image Recognition at Scale. In: International Conference on Learning Representations; 2021. Available from: https://openreview.net/forum?id=YicbFdNTTy.

27. Xie E, Wang W, Yu Z, Anandkumar A, Alvarez JM, Luo P. SegFormer: Simple and Efficient Design for Semantic Segmentation with Transformers. In: Neural Information Processing Systems (NeurIPS); 2021.

28. Hallström E. CombiANT Reader replication package; 2024. Available from: 10.5281/zenodo.13893289.

29. Harris CR, Millman KJ, van der Walt SJ, Gommers R, Virtanen P, Cournapeau D, et al. Array programming with NumPy. Nature. 2020;585(7825):357–362. doi:10.1038/s41586-020-2649-2.

30. pandas development team T. pandas-dev/pandas: Pandas; 2020. Available from: 10.5281/zenodo.3509134.

31. Paszke A, Gross S, Massa F, Lerer A, Bradbury J, Chanan G, et al. PyTorch: An Imperative Style, High-Performance Deep Learning Library. In: Advances in Neural Information Processing Systems 32. Curran Associates, Inc.; 2019. p. 8024–8035. Available from: http://papers.neurips.cc/paper/9015-pytorch-an-imperative-style-high-performance-deep-learning-library.pdf.

32. Buslaev A, Iglovikov VI, Khvedchenya E, Parinov A, Druzhinin M, Kalinin AA. Albumentations: Fast and Flexible Image Augmentations. Information. 2020;11(2). doi:10.3390/info11020125.

33. Kingma DP, Ba J. Adam: A method for stochastic optimization. arXiv preprint arXiv:14126980. 2014;.

